# Mpox virus pangenomics reveals determinants of subclade Ib

**DOI:** 10.1101/2024.10.31.24315917

**Authors:** Gustavo Sganzerla Martinez, Anuj Kumar, Eddy Kiganda-Lusamaki, Mansi Dutt, Tony Wawina-Bokalanga, Ali Toloue Ostadgavahi, Muyembe Mawete Francisca, Jean Claude Makangara-Cigolo, Patricia Kelvin, Amuri Aziza Adrienne, Christopher D Richardson, Emmanuel Lokilo, Gradi Luakanda, Ahidjo Ayouba, Anne W. Rimoin, Daniel Mukadi-Bamuleka, Eric Delaporte, Genay Pilarowski, Jason Kindrachuk, Laurens Liesenborghs, Lisa E. Hensley, Lorenzo Subissi, Martine Peeters, Nicole A. Hoff, Olivier Tshiani-Mbaya, Sofonias Tessema, Jean-Jaques Muyembe-Tamfum, Steve Ahuka-Mundeke, Alyson A Kelvin, John M Archibald, Placide Mbala-Kingebeni, Luis Flores Girón, David J Kelvin

## Abstract

Mpox, formerly monkeypox, is a viral zoonotic disease caused by the mpox virus (MPXV). MPXV, which is phylogenetically divided into Clades I and II, was declared a Public Health Emergency of International Concern for the second time in August 2024 due to rapid geographic expansion of Clade I viruses including the newly identified subclade Ib. With a unique set of genomic mutations and sustained human-to-human transmission, subclade Ib has rapidly spread throughout the eastern Democratic Republic of the Congo as well as neighboring non-endemic regions and outside the African continent. Currently, there is a lack of comparative genomic data with which to address the potential zoonotic transmissibility and pathobiology of subclade Ib. Here we report 105 protein-coding genes that are shared by all the queried MPXV subclade Ia, Ib, and IIb genomes. Our comparative genomic analysis identified that the core Clade I gene pair, *OPG032* and *OPG033*, is now a critical branching element for subclade Ia/Ib due to their loss in subclade Ib. These genes encode the complement control protein (a vaccinia virus ortholog associated with virulence), and a Kelch-like protein associated with pathogenesis, respectively, suggesting a functional evolution that might play an important role in the pathobiology of the new MPXV subclade Ib. Our results lay the groundwork to exploit the genomic elements of MPXV as potential targets for therapeutics development/repurposing, vaccine design, and molecular diagnostic expansion, as well as to uncover the viral diversity, and human-to-human transmission of MPXV.

## INTRODUCTION

Mpox virus (MPXV), the causative agent of mpox, is a double-stranded DNA poxvirus endemic to countries in Africa including the Democratic Republic of the Congo (DRC). The first human infection of MPXV was reported in the DRC in 1970 and several outbreaks with human-to-human transmission have since been reported [1]. Taxonomically, MPXV belongs to the genus *Orthopoxvirus*. MPXV can be phylogenetically divided into two clades: Clade I (formerly Congo Basin or Central African Clade) and Clade II (formerly West African Clade) [2]. The World Health Organization (WHO) declared a Public Health Emergency of International Concern (PHEIC) in 2022 as MPXV Clade IIb viruses rapidly spread to non-endemic countries and territories (n=116) where human infections and deaths were confirmed [3]. In September 2023, a new MPXV subclade Ib was first documented in Kamituga, South Kivu, DRC [4,5,6].

Subclade Ib is currently expanding to Eastern African nations [7,8] and outside the African continent with confirmed reports of cases in Germany, Sweden, Thailand, India, the United Kingdom of Great Britain and Northern Ireland, and most recently the United States and Canada. On 13 August 2024, the Africa Centres for Disease Control and Prevention (Africa CDC) declared mpox a Public Health Emergency of Continental Security [9] just before the WHO’s declaration [10] of the outbreak as a second mpox-related PHEIC in less than three years. The clinical manifestation of both mpox clades is similar [11]. Infected individuals typically develop smallpox-like symptoms including high fever, chills, headache, muscle aches, and rash that develops painful cutaneous eruptions that tend to distribute on the face, genitals, and extremities and might progress to macules, papules, vesicles, pustules, and scabs [12]. Sexual contact between individuals has been listed as a mode of transmission, but it is not exclusive; transmission by close contact with personal belongings of infected individuals has also been documented [6]. *In-vivo* data suggests the Clade Ia virus infection is more virulent with higher morbidity than the other strains [13]. Limited MPXV subclade Ib epidemiological data suggest the novel virus is more transmissible and causes lower case fatality rates when compared to previous Clade Ia outbreaks [5,6].

The large genome of MPXV encodes core genes conserved in other orthopoxviruses whose functions are involved in DNA replication, transcription, and virion assembly.

Inhibiting cytokine signaling, blocking apoptosis, and antagonizing innate immune pathways are functions commonly attributed to accessory genes found in MPXV that modulate the host immunity. The gene-rich genome of MPXV allows its adaptation to diverse hosts, highlighting the zoonotic potential and pathogenicity of MPXV [1,14].

Our objective is to derive a core genome that is shared between MPXVs subclades Ia, Ib, and IIb. Upon identifying sets of core and accessory genes that are clade-specific, we aim to generate insights that underly the transmissibility and pathogenicity of the novel subclade Ib virus. Our findings will support future efforts to (i) track MPXV viral diversity and their pathology; (ii) develop/repurpose therapeutic agents and vaccines; and (iii) create molecular diagnostic models.

## 2 MATERIALS AND METHODS

### 2.1 Public MPXV sequences

On 2025-02-05, we obtained the fasta records of MPXV sequences from the EpiPox^TM^ database hosted by the Global Initiative on Sharing All Influenza Data (GISAID). A total of 437 subclade Ia, 28 subclade Ib, and 2,627 subclade IIb sequences were downloaded as they matched the inclusion criteria of high coverage (entries with <1% Ns) and being fully sequenced (genome with > 196,000 base-pairs).

### 2.2 DNA sequencing of three MPXV subclade Ib samples

We obtained three additional MPXV subclade Ib isolates. DNA extraction of three MPXV subclade Ib samples part of the Institut National de Recherche Biomédicale (INRB) surveillance program was performed using the QIAamp® DNA Mini kit (Qiagen, Hilden, Germany). To sequence the full-length MPXV genome, libraries were made using a hybridization probes enrichment protocol with the Twist kit and the Comprehensive Viral Research Panel (Twist Biosciences). Obtained libraries were loaded onto the GridION sequencer. Consensus generation was performed as described previously [7]. Genomes are available at https://github.com/inrb-labgenpath/DRC_MPXV_Genomic_Surveillance.

### 2.3 Genome annotation

All the genomes considered in this study (n=3,092) were annotated using Prokka [15] (v1.14.5), a software tool to annotate bacterial, archaeal, and viral genomes quickly and produce standards-compliant output files. We selected clade-specific reference genomes to conduct the annotation, i.e., GenBank NC_003310.1 and NC_063383.1 for clades I and II, respectively.

### 2.4 Ortholog search

The fasta amino acids (faa) files resulting from the annotation step were used, clade-separated, as input in the OrthoFinder [16] (v2.5.5) tool to find orthologs that compose the core genomes of MPXV subclade Ia, Ib, and IIb. The resulting orthogroups files were treated in Python (v3.12) so core genes could be identified. A gene was classified as a core gene only if it was present in at least 95% of the initial poll of the genomes in each subclade. Once the orthologs present in all queried genomes were found, a set of proteins composing the core genome of each subclade was obtained.

### 2.5 Ethics approval

Samples were collected as part of national diagnostic and surveillance activities. Authorization for secondary use of the samples for research in this study was obtained from the Ethics Committee of Kinshasa School of Public Health (ESP-UNIKIN, Number ESP/CE/ 05/2023). The study samples and data were de-identified before genomic and epidemiological analyses.

## 3 RESULTS

### 3.1 MPXV clade-specific core genomes

We determined the core genomes of subclades Ia, Ib, and IIb by looking for orthologs present in at least 95% of the initial poll of the genomes of each subclade. We report having identified 134 protein-coding genes composing the core genome of MPXV subclade Ia, 148 protein-coding genes composing the core genome of subclade Ib, and 193 protein-coding genes composing the core genome of subclade IIb. We identified a total of four, two, and 30 protein-coding genes unique to the core genomes of subclade Ia, Ib, and IIb, respectively. The core genomes of subclades Ia and Ib shared two protein-coding genes, subclades Ia and IIb shared 19 protein-coding genes, and subclades Ib and IIb shared 35 protein-coding genes. In total, 105 protein-coding genes were found in common between the three subclades. In addition, we identified 4 gene duplication events in subclades Ia, Ib, and IIb. (**Figure 1**).

**Figure 1.**
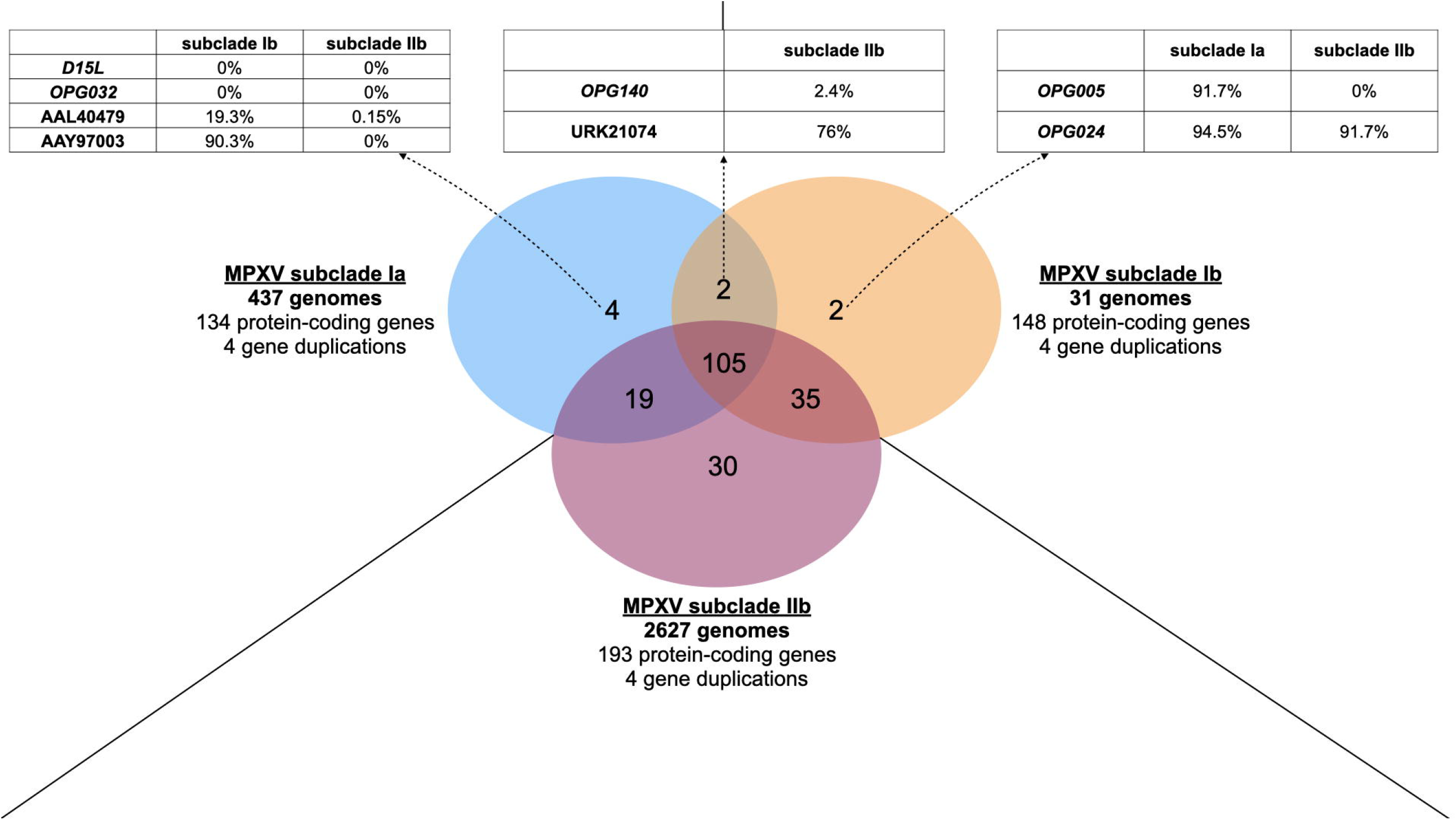
Pangenome (general MPXV) of pangenomes (subclades Ia, Ib, and IIb).

In **Figure 1**, an individual pangenome for MPXV subclade Ia, Ib, and IIb was determined. In total, 812 orthogroups were identified for subclade Ia, 309 for subclade Ib, and 243 for subclade IIb. Next, a pangenome of pangenomes was identified through a search of orthologs for the three clades, which yielded a total of 197 orthogroups.

The OrthoFinder execution of the core genomes of subclade Ia, Ib, and IIb generated a total of 197 orthogroups. Protein sequence, length, annotated function, and frequency in subclades Ia, Ib, and IIb of each orthogroup sequence is included in **Table 1**.

[Table 1]

Table 1-MPXV subclade Ia, Ib, and IIb orthogroup sequences. In **Table 1**, we include each orthogroup sequence identified in the OrthoFinder run considering the core genomes of subclade Ia, Ib, and IIb. In total, 197 orthogroups were identified. The table contains the columns i) GenBank accession number or OPG nomenclature of the protein; ii) The function of the protein per its annotation in the reference genomes of Clade I/Clade II; iii) The length of the protein in amino acid; iv) the amino acid sequence of the protein; the frequency of the protein’s presence in the subclade; v) Ia; vi) Ib; vii) IIb genomes. If a protein is indicated as being present in >95% of the genomes of a subclade, the protein composes the core genome of the subclade. ^*^ Indicates the gene in the row was found duplicated once within the subclade. ^**^ Indicates the gene in the row was found duplicated twice within the subclade. The genes duplicated are detailed in Supplementary Material S1.

### 3.2 The deletion of *OPG032* and *OPG033* genes underpins novel MPXV subclade Ib

The introduction of the novel MPXV subclade Ib featured the deletion of the *OPG032* gene [4,17]. First, we report the presence of the *OPG032* protein-coding gene in all the 437 queried MPXV clade Ia genomes. Next, we queried the annotated product referring to the *OPG032* gene in all 31 MPXV subclade Ib genomes (28 obtained from GISAID and three from this study), which was found absent in all the annotated genomes. We further confirmed the absence of the *OPG032* coding locus in subclade Ib genomes by extracting the genomic coordinates of *OPG032* in the Clade I reference genome (NC_003310.1), i.e., 19060 to 19710, and performed a nucleotide alignment of the reference genome with all 31 subclade Ib genomes (**Figure 2**). The low alignment score in the *OPG032* coordinates in all the subclade Ib genomes indicates the deletion of the *OPG032* locus in the consensus sequence of subclade Ib sequences. The genome alignments of the *OPG032* locus indicate a deletion downstream of the genomic coordinates of *OPG032*, matching the site that contains the Open Reading Frame (ORF) of the gene *OPG033*. In MPXV Clade I reference annotation, *OPG033* (19778:21291) is listed as a miscellaneous feature without a gene product directly associated to it. Despite *OPG033* not being included in the annotation file of the reference genome and consequently in the subsequent genomes annotated using it as reference, all queried MPXV subclade Ib genomes lack the DNA region that contains the *OPG033* ORF. Five random subclade Ia sequences were added in the sequence alignment for validating the excluded loci of the *OPG032* and *OPG033* gene pair in subclade Ib sequences only. In **Figure 2**, higher alignment scores are observed matching the coordinates of *OPG031* and, partially, *OPG033* in the sequences EPI_ISL_19305614, 24MPX2925V, 24MPX2832V, 24MPX3101C, EPI_ISL_19621542, and EPI_ISL_1942770. These six MPXV subclade Ib sequences were collected in 2024 in Uganda, DRC (Kinshasa), DRC (Kinshasa), DRC (Kinshasa), India (Kerala), and DRC (Kinshasa), respectively. The remaining sequences have complete deletions of *OPG032, OPG033* and partial deletions of *OPG031* and *OPG034*.

**Figure 2.**
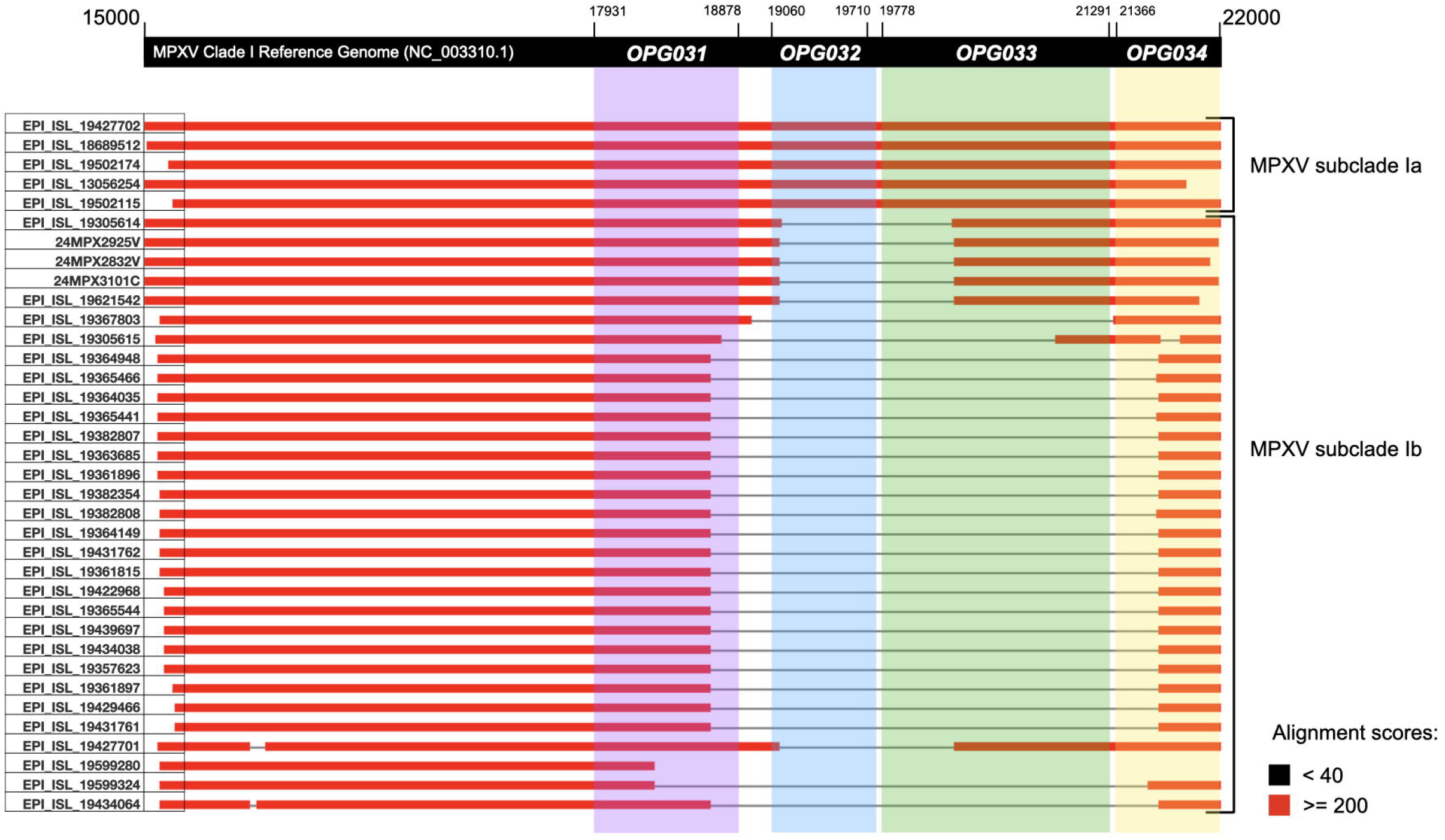
DNA alignment of deleted region in MPXV subclade Ib genomes.

**In Figure 2**, we isolated the genomic region 15000:22000 in NC_003310.1 (MPXV Clade I reference genome) which according to its annotation, contains the ORFs of *OPG031, OPG032, OPG033*, and *OPG034*. We extracted the same genomic coordinates of five random subclade Ia sequences and all 31 subclade Ib sequences analyzed in this study and aligned (blastn) the regions.

In addition to the deletion of the gene pair *OPG032* and *OPG033*, we also report the gene *OPG140* as composing the core genomes of subclade Ia and IIb; in subclade IIb, we only identified *OPG140* in 64 out of 2627 genomes queried. The gene D15L was also only located in the core genome of subclade Ia, not being present in any subclade Ib and IIb genomes. The D15L gene product is absent from the Clade I reference annotation despite its nucleotide content being present, meaning no OPG-name product is assigned to it. In other subclade Ia genomes, D15L is located at coordinates 20,197-20,514, which falls within the 19,778-21,191 region corresponding to *OPG033* in the subclade Ia reference genome, a region showing genomic erosion in subclade Ib genomes. In the subclade Ia genome (ID JX878417.1), D15L is annotated as a *provisional kelch-like protein*. The gene AAY96967 (bifunctional Toll/Il1-receptor protein) is present in the core genome of subclades Ib and IIb, with no matches found in subclade Ia genomes. The gene B15L (AAL40634) was found in the core genome of subclade Ib, 407/437 genomes of subclade Ia, and 0/2627 genomes of subclade IIb. The subclade IIb core gene *Orthopox_A49R* (AAY96964) was not found in any of the subclade Ia and Ib genomes. The subclade IIb core genes *OPG067* and QGQ59897 were found respectively in 16/437 and 415/437 subclade Ia genomes while not found in any subclade Ib genomes. Finally, the subclade IIb core gene *AAY97591* was not found in subclade Ia genomes and in 15/31 subclade Ib genomes (**Table 1**).

## 4 DISCUSSION

Our pangenome investigation allowed us to gather insights of the evolutionary trajectory of MPXVs, which showcased varying deletions and erosion in the region of the genes *OPG031*-*OPG034* and might explain the branching of Clade I into subclades a and b. Moreover, by identifying genes that are common and unique to clades and subclades of MPXVs, we deliver targets for development of therapeutics, vaccines, and molecular diagnostics methods for MPXVs from different subclades.

The 105 protein-coding genes that compose the core genome of MPXVs is similar to the 109 OPGs identified in common between different orthopoxviruses by Senkevich et al (2021) [24]. The ortholog identification in our study was conducted using OrthoFinder [16], a widely validated, general-purpose tool for comparative genomes that employs an all-versus-all sequence similarity search couple with graph-based clustering and gene tree reconstruction to infer orthogroups across complete proteomes. This strategy might be appropriate given the design of our study, which aimed to construct a pangenome based exclusively on the presence and absence of annotated protein-coding genes across different MPXV clades/subclades. Orthofinder offers methodological advantages for this purpose as it enables automated, scalable, and orthogroup clustering without requiring prior knowledge of gene families; accounts for paralogs and gene duplication events; reconstructs gene and species trees to resolve orthologous and paralogous relationships; it outputs presence/absence matrices and orthogroup composition tables that support downstream pangenome analyses, such as the identification of core and accessory sets of genes and the exploration of gene content variation among genomes. In contrast, Senkevich et al (2021) [24] employed a custom approach based on clusters of orthologs genes constructed via PSI-BLAST searches using position specific-scoring matrices derived from multiple sequence alignment of previously characterized poxvirus genes. This method is optimized for the detection of fragmented, pseudogenized, and highly divergent homologs, which favors the detailed evolutionary history of a predefined set of genes. While this approach excels in capturing cases often missed by automated workflows, it is less scalable and requires extensive manual curation on genes, making it less suitable for proteome-wide pangenome analysis where the objective is to evaluate broad patterns of gene presence and absence across genomes.

The pangenome investigation we present here provides potential targets for antiviral therapies, vaccine candidates, and molecular diagnostics. From the perspective of prevention, vaccine development efforts should consider the immunogenic as well as the conserved elements of distinct MPXV clades and subclades highlighted by our pangenome analysis. These elements could then be complemented with clade-specific features to ensure protection in cases where conserved elements alone may be insufficient. All currently used vaccines that confer protection against MPXV are vaccinia based. The smallpox vaccine was formulated with antigens of the vaccinia virus and was used to eradicate the variola virus. This vaccine confers protection across the various MPXV clades due to orthopoxviral conservation. After the eradication of smallpox, the population of vaccinated individuals in the population decreased as those born after 1980 did not routinely receive the smallpox vaccine [18]. We might expect outbreaks in naive populations with less immunity against orthopoxviruses. Proteins with antigenic properties have been found in the genome of orthopoxviruses by *in-silico, in-vitro*, and *in-vivo* methodologies [19-22]. From a treatment perspective, our data can guide therapeutic development of viral protein targets that are conserved across different MPXV variants. The protein-coding gene DNA-dependent RNA polymerase subunit 147 (*OPG105*) was identified as a core gene in of MPXV Clades I and II. The same protein was previously identified as having binding targets for inhibition with small molecules in a molecular modelling approach [23]. Taken together, the results of our core genome analysis will be of use in the development of vaccines and therapeutics against MPXV.

A major feature differentiating MPXV Clade I genomes is the presence and absence of the gene pair *OPG032* and *OPG033* in subclades Ia and Ib, respectively. The joint loss of *OPG032* and *OPG033* has previously been documented in the evolutionary history of orthopoxviruses [24]. The first reports of subclade Ib [4,17] documented the loss of the gene pair. In our work, we extended the scope of gene deletion by analyzing all available MPXV subclades Ia, Ib, and IIb genomes in a streamlined annotation pipeline. Our comparative genomic analysis enabled us to report that *OPG032* and *OPG033* were not only deleted in the novel subclade Ib but composed part of the core genome of Clade I MPXVs prior to its branching into subclades Ia and Ib. The MPXV gene *OPG032* has an ortholog in vaccinia, i.e., Vaccinia Complement Control Protein (VCP). In vaccinia, VCP is described as a virulence factor that modulates the complement system activation and inhibits early steps of the complement cascade by dissociating the C3 and C5 convertase enzymes that start and maintain the complement cascade. VCP is described as a virulence factor. Mice infected with a VCP-knockout vaccinia virus developed significantly smaller smallpox lesions than those infected with the wildtype virus [25]. An MPXV study [26] investigated the effect of incorporating and removing the *OPG032* from Clade II and I MPXV viruses, respectively. In in which prairie dogs were infected with the mutated viruses, it was observed that the removal of *OPG032* resulted in reduced mpox disease morbidity and mortality, indicating that this gene could be a significant virulence factor. However, the addition of *OPG032* into Clade II did not accelerate clinical disease course or affect disease mortality. Limited and context-dependent epidemiological data from the current outbreak of MPXV subclade Ib in eastern DRC suggests the subclade Ib viral infections in humans have lower case fatality rates than Clade Ia infections in humans [27]; however, other factors such as the socioeconomic conditions of the sites where the virus spreads, ongoing conflict in eastern DRC, and the potential presence of coinfections, reporting bias, and age of the affected population may also play a role in the sustained human-to-human transmission of MPXV subclade Ib. By highlighting a functional schism of MPXV Clade I into subclades Ia and Ib, defined by the presence or absence of the gene pair *OPG032* and *OPG033*, we lay the groundwork for questioning the impact of the complement control system in the virulence and sustained human-to-human transmission of MPXV. MPXV genomes with genomic erosion in the region of *OPG032/33* have been associated with multi-country outbreaks involving sustained human-to-human transmission. This erosion may confer a transmissibility advantage, as poxviruses are known to adapt their genomes via genomic accordions that modulate host antiviral immunity [28]. Therefore, surveillance should monitor for ne MPXV lineages with deletions in *OPG032/033* as potential proxies of sustained human-to-human transmission.

## Data Availability

All data produced in the present work are contained in the manuscript

## Acknowledgements

We appreciate the assistance provided by Dr. Nikki Kelvin for input regarding poxviruses. We also acknowledge participation of the Digital Research Alliance of Canada through its Atlantic Canadian provider ACENET for providing high-performance computing infrastructure to partially run the analyses depicted in this work. All the MPXV genome sequences were obtained from the Global Initiative on Sharing All Influenza Data (GISAID). We acknowledge the laboratories in which the MPXVs were originally obtained. The authors of this work, Authorized Users, relinquish any ownership rights of the intellectual properties deposited by the following laboratories. The detailed mentioning of the laboratories, per GISAID data sharing policies, is included in Supplementary Material S2.

