## Supplementary Material S1 for "Mpox virus pangenomics reveals determinants of subclade Ib"

The OrthoFinder execution of the pangenome (MPXV) of pangenomes (MPXV subclade Ia, Ib, and IIb) resulted in 197 orthogroups. Gene duplication events were in six orthogroups.

1. OG0000005 (serpin) was duplicated in subclade Ia sequences (*OPG199* and *OPG208*).
2. OG0000001 (C4L/C10L-like family protein) was duplicated twice in subclade IIb sequences (*OPG031*, QGQ59729, and QGQ59730).
3. OG0000002 (Ankyrin repeat protein [44]) was duplicated in subclade Ib and IIb sequences (*OPG003* and *OPG205*).
4. OG0000003 (Phospholipase-D-like protein) was duplicated in subclade Ia and Ib sequences (*OPG042* and *OPG057*).
5. OG0000004 (Ankyrin-like protein [1]) was duplicated in subclade Ia and IIb sequences (*OPG037* and *OPG189*).
6. OG0000000 (Ankyrin repeat protein [14]) was duplicated in subclade Ia and duplicated twice in subclade Ib (*OPG025*, *OPG023*, and *OPG015*).
