## Supplementary Material S1 for "Mpox virus pangenomics reveals determinants of subclade Ib"

The Monkeypox virus (MPXV) genomes used in this work were obtained from the EpiPoxTM database hosted by the Global Initiative on Sharing All Influenza Data (GISAID) on 2025-02-05. We acknowledge the following laboratories in which the MPXVs were originally obtained. The authors of this work, Authorized Users, relinquish any ownership rights of the intellectual properties deposited by the following laboratories.

Área de Innovación y Desarrollo. Centro Nacional de Salud Publica. Instituto Nacional de Salud, Laboratorio de Referencia Nacional de Virus Inmunoprevenibles. Centro Nacional de Salud Publica. Instituto Nacional de Salud, South America / Peru / Lima / Lima / Santiago De Surco

Laboratorio de Referencia Nacional de Virus Respiratorio. Centro Nacional de Salud Publica. Instituto Nacional de Salud., Laboratorio de Referencia Nacional de Virus Respiratorio. Centro Nacional de Salud Publica. Instituto Nacional de Salud., South America / Peru / Loreto

Instituto Adolfo Lutz, Rapid Response Center, Strategic Laboratory, Vigilancia Epidemiologica, South America / Brazil / Sao Paulo / Franca

Instituto Adolfo Lutz Strategic Laboratory, USF Boicucanga I Sao Sebastiao, South America / Brazil / Sao Paulo / Sao Sebastiao

Laboratorio Nacional de Salud Pública Dr. Defilló, Laboratorio Nacional de Salud Pública Dr. Defilló, North America / Dominican Republic / Distrito Nacional

CDCT/CEVS/SES-RS, CDCT/CEVS/SES-RS, South America / Brazil / Rio Grande do Sul / Parobe

Laboratory of Virology, University Hospitals of Geneva, Laboratory of Virology, University Hospitals of Geneva, Europe / Switzerland

Instituto de Diagnostico y Referencia Epidemiologicos (INDRE), LESP Chihuahua, North America / Mexico / Chihuahua

Bioinformatic lab, Scientific Institute IRCCS E. Medea, Sexually Transmitted Diseases (STDs) outpatient service of Dermatology Unit, Fondazione IRCCS Ca' Granda Ospedale Maggiore Policlinico of Milan, Europe / Italy

Laboratorio Nacional de Salud Pública Dr. Defilló, Laboratorio Nacional de Salud Pública Dr. Defilló, North America / Dominican Republic / La Romana

Environmental, Agricultural, and Occupational Health, University of Nebraska Medical Center, Environmental, Agricultural, and Occupational Health, University of Nebraska Medical Center, North America / USA / Iowa

Instituto de Salud Publica de Chile, Genetica Molecular and Subdepartamento de Virologia ISP Chile, South America / Chile / Region Metropolitana de Santiago / Quinta Normal

Balai Besar Laboratorium Biologi Kesehatan, PKC Tanjung Priok, Asia / Indonesia / DKI Jakarta / Jakarta Utara

Centro de Desenvolvimento Científico e Tecnológico (CDCT), Centro Estadual de Vigilância em Saúde (CEVS) da Secretaria Estadual da Saúde (SES-RS), Centro de Desenvolvimento Científico e Tecnológico (CDCT), Centro Estadual de Vigilância em Saúde (CEVS) da Secretaria Estadual da Saúde (SES-RS), South America / Brazil / Rio Grande do Sul / Campo Bom

Instituto Nacional de Saude Doutor Ricardo Jorge (INSA), Instituto Nacional de Saude Doutor Ricardo Jorge (INSA), Europe / Portugal

Department of Medical Microbiology & Infection prevention, Amsterdam University Medical Centers location AMC, Department of Medical Microbiology & Infection prevention, Amsterdam University Medical Centers location AMC, Europe / Netherlands / Noord-Holland / Amsterdam

Instituto Adolfo Lutz Strategic Laboratory, UPA Vila Mariana, South America / Brazil / Sao Paulo / Sao Paulo

Microbiology Department. Complexo Hospitalario Universitario de Vigo, Complejo Hospitalario Universitario de Pontevedra, Europe / Spain / Galicia / Vigo

Public Health Agency of Canada, National Microbiology Laboratory, Public Health Agency of Canada, National Microbiology Laboratory, North America / Canada

Pathogen Genomic Laboratory, Institut National de Recherche Biomedicale (INRB), Pathogen Genomic Laboratory, Institut National de Recherche Biomedicale (INRB), Africa / Democratic Republic of the Congo / Equateur / Mbandaka

Instituto Adolfo Lutz, Rapid Response Center, Strategic Laboratory, Unidade Basica de Saude Chacara Cruzeiro do Sul, South America / Brazil / Sao Paulo / Sao Paulo

Genomics Division, Instituto Tecnologico y de Energias Renovables (ITER), Poligono Industrial de Granadilla,, Genomics Division, Instituto Tecnologico y de Energias Renovables (ITER), Poligono Industrial de Granadilla, Europe / Spain

Alberta Precision Laboratories, Alberta Precision Laboratories, North America / Canada / Alberta

Public and Environmental Health Reference Laboratories, Pathology Queensland, Queensland Medical Laboratories, Oceania / Australia / Queensland

National Institute of Health Research and Development, PKC Kramat Jati, Asia / Indonesia / DKI Jakarta / Jakarta Timur

Instituto de Salud Publica de Chile, Genetica Molecular and Subdepartamento de Virologia ISP Chile, South America / Chile / Valparaiso / Valparaiso

Laboratorio de Referencia Nacional de Virus Inmunoprevenibles. Centro Nacional de Salud Publica. Instituto Nacional de Salud., Laboratorio de Referencia Nacional de Virus Inmunoprevenibles. Centro Nacional de Salud Publica. Instituto Nacional de Salud., South America / Peru / Lambayeque

Instituto Adolfo Lutz, Rapid Response Center, Strategic Laboratory, Hospital Santa Catarina, South America / Brazil / Sao Paulo / Sao Paulo

Pathogen Genomic Laboratory, Institut National de Recherche Biomedicale (INRB), Pathogen Genomic Laboratory, Institut National de Recherche Biomedicale (INRB), Africa / Democratic Republic of the Congo / Tshopo

Inciensa, Investigación y Enseñanza en Nutrición y Salud Centro Nacional de Referencia de Virología, Division de Microbiologia, Hospital Nacional de Niños Carlos Saenz Herrera, North America / Costa Rica / San Jose

KU Leuven, Rega Institute, Department of Microbiology, Immunology and Transplantation, KU Leuven, Rega Institute, Department of Microbiology, Immunology and Transplantation, Europe / Belgium

Instituto Adolfo Lutz Strategic Laboratory, Hospital Santa Ignes, South America / Brazil / Sao Paulo / Indaiatuba

Laboratorio de Referencia Nacional de Virus Respiratorio. Centro Nacional de Salud Publica. Instituto Nacional de Salud., Laboratorio de Referencia Nacional de Virus Respiratorio. Centro Nacional de Salud Publica. Instituto Nacional de Salud., South America / Peru / Tacna

Viral and Rickettsial Disease Laboratory (VRDL) California Department of Public Health (CDPH), Viral and Rickettsial Disease Laboratory (VRDL) California Department of Public Health (CDPH), North America / USA / California

Instituto Adolfo Lutz Strategic Laboratory, Ambulatorio de Atendimentode DST de Guariba, South America / Brazil / Sao Paulo / Guariba

Instituto de Diagnostico y Referencia Epidemiologicos (INDRE), LESP San Luis Potosi, North America / Mexico / San Luis Potosi

Biochemistry and Microbiology, University of Victoria, Biochemistry and Microbiology, University of Victoria, Africa / Sudan / Nuri

Instituto Adolfo Lutz Strategic Laboratory, CTA Centro de Testagem e Aconselhamento Favo de Mel, South America / Brazil / Sao Paulo / Jandira

Laboratorio de Referencia Nacional de Virus Respiratorio. Centro Nacional de Salud Publica. Instituto Nacional de Salud., Laboratorio de Referencia Nacional de Virus Respiratorio. Centro Nacional de Salud Publica. Instituto Nacional de Salud., South America / Peru / Piura

Laboratorio de Referencia Nacional de Viruas Inmunoprevenibles. Centro Nacional de Salud Publica. Instiuto Nacional de Salud, Laboratorio de Referencia Nacional de Viruas Inmunoprevenibles. Centro Nacional de Salud Publica. Instiuto Nacional de Salud, South America / Peru / Lima / Lima / Ate Vitarte

Laboratory of Genomics and Bioinformatics, Comenius University Science Park, Public Health Authority of the Slovak Republic, Europe / Slovakia / Brezno

Bioinformatic Lab, Scientific Institute IRCCS E. Medea, Sexually Transmitted Diseases (STDs) outpatient service of Dermatology Unit, Fondazione IRCCS Ca' Granda Ospedale Maggiore Policlinico of Milan, Europe / Italy

NC - Division of High-Consequence Pathogens and Pathology (DHCPP)- Poxvirus and Rabies Branch (PRB), NC - Division of High-Consequence Pathogens and Pathology (DHCPP)- Poxvirus and Rabies Branch (PRB), Africa / Cameroon

Instituto Adolfo Lutz, Rapid Response Center, Strategic Laboratory, Ubs Alexander Fleming Simioni, South America / Brazil / Sao Paulo / Ribeirao Preto

CDCT/CEVS/SES-RS, CDCT/CEVS/SES-RS, South America / Brazil / Rio Grande do Sul / Estancia Velha

Instituto Adolfo Lutz Strategic Laboratory, UBS Jardim Santista, South America / Brazil / Sao Paulo / Maua

Pathogen Genomic Laboratory, Institut National de Recherche Biomedicale (INRB), Pathogen Genomic Laboratory, Institut National de Recherche Biomedicale (INRB), Africa / Democratic Republic of the Congo / Maniema

Laboratory Medicine and Pathology, University of Washington, Laboratory Medicine and Pathology, University of Washington, North America / USA

Robert Koch Institute, Centre for Biological Threats, Highly Pathogenic Viruses, Robert Koch Institute, Centre for Biological Threats, Highly Pathogenic Viruses, Europe / Germany

Instituto de Salud Publica de Chile, Genetica Molecular and Subdepartamento de Virologia ISP Chile, South America / Chile / Coquimbo / La Serena

Balai Besar Laboratorium Biologi Kesehatan, Eka Hospital BSD, Asia / Indonesia / Banten / Tangerang Selatan

Instituto Adolfo Lutz Strategic Laboratory, SAE DST AIDS M Boi Mirim Servico de Atencao Especializada, South America / Brazil / Sao Paulo / Sao Paulo

Laboratorio de Referencia Nacional de Viruas Inmunoprevenibles. Centro Nacional de Salud Publica. Instiuto Nacional de Salud, Laboratorio de Referencia Nacional de Viruas Inmunoprevenibles. Centro Nacional de Salud Publica. Instiuto Nacional de Salud, South America / Peru / Arequipa / Arequipa / Arequipa

Direccion de Investigacion en Salud Publica, Instituto Nacional de Salud, Direccion de Investigacion en Salud Publica, Instituto Nacional de Salud, South America / Colombia / Valle del Cauca

Instituto Adolfo Lutz Strategic Laboratory, AMA Capao Redondo, South America / Brazil / Sao Paulo / Sao Paulo

Instituto Adolfo Lutz, Rapid Response Center, Strategic Laboratory, Instituto Adolfo Lutz, Rapid Response Center, Strategic Laboratory, South America / Brazil / Sao Paulo / Tiete

Centro de Desenvolvimento Científico e Tecnológico (CDCT), Centro Estadual de Vigilância em Saúde (CEVS) da Secretaria Estadual da Saúde (SES-RS), Centro de Desenvolvimento Científico e Tecnológico (CDCT), Centro Estadual de Vigilância em Saúde (CEVS) da Secretaria Estadual da Saúde (SES-RS), South America / Brazil / Rio Grande do Sul / Viamao

Laboratorio de Referencia Nacional de Viruas Inmunoprevenibles. Centro Nacional de Salud Publica. Instiuto Nacional de Salud, Laboratorio de Referencia Nacional de Viruas Inmunoprevenibles. Centro Nacional de Salud Publica. Instiuto Nacional de Salud, South America / Peru / La Libertad / Trujillo / Huanchaco

Instituto de Salud Publica de Chile, Genetica Molecular and Subdepartamento de Virologia ISP Chile, South America / Chile / Region Metropolitana de Santiago / Puente Alto

Instituto Adolfo Lutz, Rapid Response Center, Strategic Laboratory, Instituto Adolfo Lutz, Rapid Response Center, Strategic Laboratory, South America / Brazil / Sao Paulo / Barueri

Instituto Adolfo Lutz Strategic Laboratory, Hosp. Carlos Chagas, South America / Brazil / Sao Paulo / Guarulhos

Istituto Zooprofilattico Sperimentale della Puglia e della Basilicata, Laboratorio di Epidemiologia Molecolare e Sanità Pubblica-Policlinico Bari, Europe / Italy / Bari

Pathogen Genomic Laboratory, Institut National de Recherche Biomedicale, Pathogen Genomic Laboratory, Institut National de Recherche Biomedicale, Africa / Democratic Republic of the Congo

Instituto de Diagnostico y Referencia Epidemiologicos (INDRE), LESP Coahuila, North America / Mexico / Coahuila

Laboratorio de Referencia Nacional de Virus Inmunoprevenibles. Centro Nacional de Salud Publica. Instituto Nacional de Salud., Laboratorio de Referencia Nacional de Virus Inmunoprevenibles. Centro Nacional de Salud Publica. Instituto Nacional de Salud., South America / Peru / Huanuco

Instituto Oswaldo Cruz FIOCRUZ - Laboratory of Respiratory Viruses and Measles (LVRS), Laboratorio de Enterovirus, Instituto Oswaldo Cruz, Fiocruz, South America / Brazil / Rio de Janeiro / Rio de Janeiro

Cellule d'Intervention Biologique d'Urgence, Institut Pasteur, Cellule d'Intervention Biologique d'Urgence, Institut Pasteur, Europe / France

Virologia, INEI- ANLIS Dr. Carlos G. Malbran, Virologia, INEI- ANLIS Dr. Carlos G. Malbran, South America / Argentina

Instituto Adolfo Lutz Strategic Laboratory, Pronto Socorro Municiplal de Cravinhos, South America / Brazil / Sao Paulo / Cravinhos

National Institute of Health Research and Development, RS Brawijaya Saharjo, Asia / Indonesia / DKI Jakarta / Jakarta Selatan

Laboratory of Genomics and Bioinformatics, Comenius University Science Park, Public Health Authority of the Slovak Republic, Europe / Slovakia / Bratislava

Pathogen Genomics Lab, National Institute for Biomedical Research (INRB), Pathogen Genomics Lab, National Institute for Biomedical Research (INRB), Africa / Democratic Republic of the Congo / Kinshasa

Environmental, Agricultural, and Occupational Health, University of Nebraska Medical Center, Nebraska Public Health Laboratory (NPHL), North America / USA / Oklahoma

Genomics and Epigenomics, AREA Science Park, SC (UCO) Igiene e Sanità Pubblica, ASUGI, Trieste, Europe / Italy / Gorizia

Microbiology, Immunology and Transplantation, KU Leuven, Rega Institute, Microbiology, Immunology and Transplantation, KU Leuven, Rega Institute, Europe / Belgium

Division of High-risk Pathogens, Korea Disease Control and Prevention Agency, Division of High-risk Pathogens, Korea Disease Control and Prevention Agency, Asia / South Korea

CDCT/CEVS/SES-RS, CDCT/CEVS/SES-RS, South America / Brazil / Rio Grande do Sul / Novo Hamburgo

Instituto Adolfo Lutz Strategic Laboratory, Secretaria Municipal da Saude de Joanopolis, South America / Brazil / Sao Paulo / Joanopolia

Medical University of Vienna Center for Virology, Center for Virology, Medical University of Vienna, Europe / Austria / Vorarlberg

Laboratorio de Referencia Nacional de Viruas Inmunoprevenibles. Centro Nacional de Salud Publica. Instiuto Nacional de Salud, Laboratorio de Referencia Nacional de Viruas Inmunoprevenibles. Centro Nacional de Salud Publica. Instiuto Nacional de Salud, South America / Peru / Lima / Lima / Independencia

Laboratorio de Referencia Nacional de Viruas Inmunoprevenibles. Centro Nacional de Salud Publica. Instiuto Nacional de Salud, Laboratorio de Referencia Nacional de Viruas Inmunoprevenibles. Centro Nacional de Salud Publica. Instiuto Nacional de Salud, South America / Peru / Callao

Laboratorio de Referencia Nacional de Viruas Inmunoprevenibles. Centro Nacional de Salud Publica. Instiuto Nacional de Salud, Laboratorio de Referencia Nacional de Viruas Inmunoprevenibles. Centro Nacional de Salud Publica. Instiuto Nacional de Salud, South America / Peru / La Libertad / Trujillo / Florencia de Mora

National Institute of Health Research and Development, PKM Pancoran, Asia / Indonesia / DKI Jakarta / Jakarta Selatan

Medical University of Vienna Center for Virology, Center for Virology Medical Unviersity of Vienna, Europe / Austria / Lower Austria

Instituto Adolfo Lutz, Rapid Response Center, Strategic Laboratory, Hospital das Clinicas da Unicamp De Campinas, South America / Brazil / Sao Paulo / Campinas

Regional Innovative Public Health Laboratory (RIPHL) at Rush University Medical Center, Quest Diagnostics, North America / USA / Illinois / Cook County / Chicago

Laboratorio de Referencia Nacional de Viruas Inmunoprevenibles. Centro Nacional de Salud Publica. Instiuto Nacional de Salud, Laboratorio de Referencia Nacional de Viruas Inmunoprevenibles. Centro Nacional de Salud Publica. Instiuto Nacional de Salud, South America / Peru / Lambayeque / Chiclayo / Chiclayo

Pathogen Genomic Laboratory, Institut National de Recherche Biomedicale (INRB), Pathogen Genomic Laboratory, Institut National de Recherche Biomedicale (INRB), Africa / Democratic Republic of the Congo / Mai-Ndombe / Inongo

National Public Health Center, National Biosafety Laboratory, National Public Health Center, National Biosafety Laboratory, Europe / Hungary / Budapest

Laboratorio de Referencia Nacional de Viruas Inmunoprevenibles. Centro Nacional de Salud Publica. Instiuto Nacional de Salud, Laboratorio de Referencia Nacional de Viruas Inmunoprevenibles. Centro Nacional de Salud Publica. Instiuto Nacional de Salud, South America / Peru / Loreto / Maynas / Punchana

Instituto Adolfo Lutz, Rapid Response Center, Strategic Laboratory, Ambulatorio de Molestias Infectocontagiosas, South America / Brazil / Sao Paulo / Jundiai

USAMRIID, Center for Genome Sciences, United States Army Medical Research Institute of Infectious Diseases, USAMRIID, Center for Genome Sciences, United States Army Medical Research Institute of Infectious Diseases, Africa / Democratic Republic of the Congo

Instituto Adolfo Lutz Strategic Laboratory, UBS Horto Florestal, South America / Brazil / Sao Paulo / Sao Paulo

Instituto Adolfo Lutz Strategic Laboratory, Centro de Referencia Modulo I SAE II Bauru, South America / Brazil / Sao Paulo / Bauru

Institute of Ecology and Evolution, University of Edinburgh, Nigeria Centre for Disease Control and Prevention, Africa / Nigeria

Instituto Adolfo Lutz Strategic Laboratory, Secretaria de Saude de Mogi das Cruzes, South America / Brazil / Sao Paulo / Mogi das Cruzes

Laboratorio de Referencia Nacional de Viruas Inmunoprevenibles. Centro Nacional de Salud Publica. Instiuto Nacional de Salud, Laboratorio de Referencia Nacional de Viruas Inmunoprevenibles. Centro Nacional de Salud Publica. Instiuto Nacional de Salud, South America / Peru / Lima / Lima / Villa El Salvador

Hospital Universitari Vall d'Hebron, Hospital Universitari Vall d'Hebron, Europe / Spain / Catalunya

Laboratorio de Referencia Nacional de Virus Respiratorio. Centro Nacional de Salud Publica. Instituto Nacional de Salud., Laboratorio de Referencia Nacional de Virus Respiratorio. Centro Nacional de Salud Publica. Instituto Nacional de Salud., South America / Peru / Lima

Instituto Adolfo Lutz, Rapid Response Center, Strategic Laboratory, Instituto Adolfo Lutz, Rapid Response Center, Strategic Laboratory, South America / Brazil / Sao Paulo / Caraguatatuba

National Institute of Health Research and Development, PKC Pancoran, Asia / Indonesia / DKI Jakarta / Jakarta Selatan

Instituto Adolfo Lutz Strategic Laboratory, Hosp. Alemao Oswaldo Cruz, South America / Brazil / Sao Paulo / Sao Bernado do Campo

Centro de Desenvolvimento Científico e Tecnológico (CDCT), Centro Estadual de Vigilância em Saúde (CEVS) da Secretaria Estadual da Saúde (SES-RS), Centro de Desenvolvimento Científico e Tecnológico (CDCT), Centro Estadual de Vigilância em Saúde (CEVS) da Secretaria Estadual da Saúde (SES-RS), South America / Brazil / Rio Grande do Sul / Passo Fundo

Instituto Adolfo Lutz Strategic Laboratory, Pronto Atendimento Infantil e Central de Quimioterapia de Sao Jose do Rio Preto, South America / Brazil / Sao Paulo / Sao Jose do Rio Preto

Instituto Adolfo Lutz, Rapid Response Center, Strategic Laboratory, Centro de Diagnostico Secedi Santos, South America / Brazil / Sao Paulo / Santos

National Virus Reference Laboratory, National Virus Reference Laboratory, Europe / Ireland / Cork

División Diagnóstico Molecular Hospital México, División Diagnóstico Molecular Hospital México, North America / Costa Rica / San Jose

Instituto de Salud Publica de Chile, Genetica Molecular and Subdepartamento de Virologia ISP Chile, South America / Chile / Region Metropolitana de Santiago / Talagante

Instituto de Diagnostico y Referencia Epidemiologicos (INDRE), LESP Chiapas, North America / Mexico / Chiapas

Instituto Adolfo Lutz Strategic Laboratory, Centro de Saude 24 horas, South America / Brazil / Sao Paulo / Itaquaquecetuba

Pathogen Genomic Laboratory, Institut National de Recherche Biomedicale (INRB), Pathogen Genomic Laboratory, Institut National de Recherche Biomedicale (INRB), Africa / Democratic Republic of the Congo

Department of Medical Microbiology & Infection prevention, Amsterdam University Medical Centers location AMC, Public Health Laboratory, Public Health Service Amsterdam, The Netherlands, Europe / Netherlands / North-Holland / Amsterdam

Jiangsu Centers for Diseases Prevention and Control, Jiangsu Centers for Diseases Prevention and Control, Asia / China / Jiangsu / Nantong

Instituto de Diagnostico y Referencia Epidemiologicos (INDRE), LESP Guerrero, North America / Mexico / Guerrero

Instituto Adolfo Lutz Strategic Laboratory, UBS Parque Meia Lua, South America / Brazil / Sao Paulo / Jacarei

Oswaldo Cruz Foundation Laboratory of Respiratory Virus and Measles, Laboratorio de Enterovirus, Instituto Oswaldo Cruz, Fiocruz, South America / Brazil / Rio de Janeiro / Rio de Janeiro

Genomics Division, Instituto Tecnologico y de Energias Renovables (ITER), Genomics Division, Instituto Tecnologico y de Energias Renovables (ITER), Europe / Spain

Instituto Adolfo Lutz Strategic Laboratory, Instituto de Infectologia Emilio Ribas, South America / Brazil / Sao Paulo / Sao Paulo

National Institute for Communicable Diseases of the National Health Laboratory Service, National Institute for Communicable Diseases of the National Health Laboratory Service, Africa / South Africa / Western Cape

Instituto Adolfo Lutz Strategic Laboratory, Pronto Socorro da Vila Dirce, South America / Brazil / Sao Paulo / Carapicuiba

Biochemistry and Molecular Genetics, Israel Institute for Biological Research, Biochemistry and Molecular Genetics, Israel Institute for Biological Research, Asia / Israel

Laboratorio de Referencia Nacional de Virus Respiratorios. Centro Nacional de Salud Publica. Instituto Nacional de Salud Peru., Laboratorio de Referencia Nacional de Virus Respiratorios. Centro Nacional de Salud Publica. Instituto Nacional de Salud Peru., South America / Peru / Lima

Instituto Adolfo Lutz, Rapid Response Center, Strategic Laboratory, Unidade Mista de Atendimento Infantil de Carapicuiba, South America / Brazil / Sao Paulo / Carapicuiba

Laboratorio de Referencia Nacional de Virus Inmunoprevenibles. Centro Nacional de Salud Publica. Instituto Nacional de Salud., Laboratorio de Referencia Nacional de Virus Inmunoprevenibles. Centro Nacional de Salud Publica. Instituto Nacional de Salud., South America / Peru / Arequipa

University of Rochester Medical Center, University of Rochester Medical Center, North America / USA / New York

Instituto de Salud Publica de Chile, Genetica Molecular and Subdepartamento de Virologia ISP Chile, South America / Chile / Biobio / Concepcion

Instituto Adolfo Lutz Strategic Laboratory, Hospital Nossa Senhora de Lourdes, South America / Brazil / Sao Paulo / Sao Paulo

Cellule d'Intervention Biologique d'Urgence, Institut Pasteur, Centre Médical de l'Institut Pasteur, Europe / France

Instituto Adolfo Lutz, Rapid Response Center, Strategic Laboratory, Instituto Adolfo Lutz, Rapid Response Center, Strategic Laboratory, South America / Brazil / Sao Paulo / Guarulhos

Instituto de Diagnostico y Referencia Epidemiologicos (INDRE), LESP Zacatecas, North America / Mexico / Zacatecas

Victorian Infectious Diseases Reference Laboratory (VIDRL), Melbourne Health, Victorian Infectious Diseases Reference Laboratory (VIDRL), Melbourne Health, Oceania / Australia / Victoria

Instituto de Salud Publica de Chile, Genetica Molecular and Subdepartamento de Virologia ISP Chile, South America / Chile / Region Metropolitana de Santiago / Las Condes

Division of Infectious Disease Vaccine Research, Korea National Institute of Health, Institut National de Recherche Biomedicale, Africa / Democratic Republic of the Congo

Laboratorio de Referencia Nacional de Viruas Inmunoprevenibles. Centro Nacional de Salud Publica. Instiuto Nacional de Salud, Laboratorio de Referencia Nacional de Viruas Inmunoprevenibles. Centro Nacional de Salud Publica. Instiuto Nacional de Salud, South America / Peru / Lima / Lima / San Martin de Porres

Instituto de Diagnostico y Referencia Epidemiologicos (INDRE), LESP Queretaro, North America / Mexico / Queretaro

Instituto Adolfo Lutz Strategic Laboratory, Centro de Referencia em Especialidades Central Rib Preto, South America / Brazil / Sao Paulo / Ribeirao Preto

Microbiology Service, Hospital Universitario Clinico San Cecilio, Granada, Microbiology Service, Hospital Universitario Clinico San Cecilio, Granada, Europe / Spain / Andalusia

University of Bologna, Department of Medical and Surgical Sciences (DIMEC), University of Bologna, Department of Medical and Surgical Sciences (DIMEC), Europe / Italy / Lomagna

Pathogen Genomic Laboratory, Institut National de Recherche Biomedicale (INRB), Pathogen Genomic Laboratory, Institut National de Recherche Biomedicale (INRB), Africa / Democratic Republic of the Congo / Maniema / Kindu

Instituto de Salud Publica de Chile, Genetica Molecular and Subdepartamento de Virologia ISP Chile, South America / Chile / Valparaiso / San Antonio

Instituto Adolfo Lutz Strategic Laboratory, Pronto Socorro Municipal do Promorar, South America / Brazil / Piaui / Teresina

Instituto Adolfo Lutz, Rapid Response Center, Strategic Laboratory, Instituto Adolfo Lutz, Rapid Response Center, Strategic Laboratory, South America / Brazil / Sao Paulo / Valinhos

Laboratorio de Referencia Nacional de Viruas Inmunoprevenibles. Centro Nacional de Salud Publica. Instiuto Nacional de Salud, Laboratorio de Referencia Nacional de Viruas Inmunoprevenibles. Centro Nacional de Salud Publica. Instiuto Nacional de Salud, South America / Peru / Lima

Instituto Adolfo Lutz, Rapid Response Center, Strategic Laboratory, Instituto Adolfo Lutz, Rapid Response Center, Strategic Laboratory, South America / Brazil / Sao Paulo / Ribeirao Preto

Instituto de Salud Publica de Chile, Genetica Molecular and Subdepartamento de Virologia ISP Chile, South America / Chile / Region Metropolitana de Santiago / San Miguel

Instituto de Diagnostico y Referencia Epidemiologicos (INDRE), LESP Oaxaca, North America / Mexico / Oaxaca

Biochemistry and Microbiology, University of Victoria, Biochemistry and Microbiology, University of Victoria, Africa / Democratic Republic of the Congo

Department of Virology, National Institute of Health, Public Health Reference Laboratory, Khyber Medical University, Asia / Pakistan / Peshawar

Instituto de Diagnostico y Referencia Epidemiologicos (INDRE), LESP Baja California, North America / Mexico / Baja California

IRBA Research Institute Biomédicale Des Armées, IRBA Research Institute Biomédicale Des Armées, Europe / France

Instituto Adolfo Lutz, Rapid Response Center, Strategic Laboratory, Hospital San Paolo Santana, South America / Brazil / Sao Paulo / Sao Paulo

Laboratorio de Referencia Nacional de Viruas Inmunoprevenibles. Centro Nacional de Salud Publica. Instiuto Nacional de Salud, Laboratorio de Referencia Nacional de Viruas Inmunoprevenibles. Centro Nacional de Salud Publica. Instiuto Nacional de Salud, South America / Peru / Ica / Ica / Ica

Instituto Adolfo Lutz Strategic Laboratory, Hospital e Maternidade Santa Maria Cruz Azul, South America / Brazil / Sao Paulo / Sao Paulo

Pathogen Genomic Laboratory, Institut National de Recherche Biomedicale (INRB), Pathogen Genomic Laboratory, Institut National de Recherche Biomedicale (INRB), Africa / Democratic Republic of the Congo / Mongala

Instituto Adolfo Lutz, Rapid Response Center, Strategic Laboratory, Hospital Renascenca Campinas, South America / Brazil / Sao Paulo / Campinas

Laboratory Medicine and Pathology, University of Washington, Laboratory Medicine and Pathology, University of Washington, North America / USA / California

Thai Red Cross Emerging Infectious Diseases Clinical Center and Faculty of Medicine, Chulalongkorn University, Vajira Hospital, Asia / Thailand

National Institute of Health Research and Development, RSUPN dr. Cipto Mangunkusumo, Asia / Indonesia / Jakarta

Instituto Adolfo Lutz, Rapid Response Center, Strategic Laboratory, Upa Atalaia Dra Rita De Cassia Sorio, South America / Brazil / Sao Paulo / Cotia

Institute of Medical Virology, University of Zurich, checkin Zollhaus, Europe / Switzerland / Zurich

Institute of Ecology and Evolution, University of Edinburgh, Nigeria Centre for Disease Control and Prevention, Africa / Nigeria / Cross River

National Institute of Health Research and Development, PKC Kebayoran Lama, Asia / Indonesia / DKI Jakarta / Jakarta Selatan

Microbiol Genomics and Bioinformatics, Bundeswehr Institute of Microbiology, Microbiol Genomics and Bioinformatics, Bundeswehr Institute of Microbiology, Europe / Germany / Bavaria

National Institute of Health Research and Development, PKC Grogol Petamburan, Asia / Indonesia / DKI Jakarta / Jakarta Barat

Instituto Adolfo Lutz, Rapid Response Center, Strategic Laboratory, Hospital Santa Marcelina, South America / Brazil / Sao Paulo / Sao Paulo

Instituto Adolfo Lutz, Rapid Response Center, Strategic Laboratory, Centro de Doencas Infecto Contagiosas Cedic de Piracicaba, South America / Brazil / Sao Paulo / Piracicaba

Department of Infectious Diseases, National Institute of Health Doutor Ricardo Jorge, Portugal (INSA), Department of Infectious Diseases, National Institute of Health Doutor Ricardo Jorge, Portugal (INSA), Europe / Portugal

Environmental, Agricultural, and Occupational Health, University of Nebraska Medical Center, 984388 Nebraska Medical Center, Environmental, Agricultural, and Occupational Health, University of Nebraska Medical Center, 984388 Nebraska Medical Center, North America / USA / Nebraska

Pathogen Genomic Laboratory, Institut National de Recherche Biomedicale (INRB), Pathogen Genomic Laboratory, Institut National de Recherche Biomedicale (INRB), Africa / Democratic Republic of the Congo / KasaÃ¯

Viral and Rickettsial Disease Laboratory, California Department of Public Health, Viral and Rickettsial Disease Laboratory, California Department of Public Health, North America / USA / California

Instituto Adolfo Lutz Strategic Laboratory, UBS Alexander Fleming Simioni, South America / Brazil / Sao Paulo / Ribeirao Preto

Los Angeles County Public Health Laboratories, Kaiser Permanente Chino Hills Regional Reference Laboratories, North America / USA / California / Los Angeles County

National Institute of Health Research and Development, PKC Senen, Asia / Indonesia / DKI Jakarta / Jakarta Pusat

Public Health Laboratory, NYC Department of Health and Mental Hygiene (DOHMH), Public Health Laboratory, NYC Department of Health and Mental Hygiene (DOHMH), North America / USA / New York / New York City

Laboratorio de Referencia Nacional de Viruas Inmunoprevenibles. Centro Nacional de Salud Publica. Instiuto Nacional de Salud, Laboratorio de Referencia Nacional de Viruas Inmunoprevenibles. Centro Nacional de Salud Publica. Instiuto Nacional de Salud, South America / Peru / Lima / Lima / Breña

IRCCS Sacro Cuore Don Calabria Hospital, Department of Infectious, Tropical Diseases & Microbiology, IRCCS Sacro Cuore Don Calabria Hospital, Department of Infectious, Tropical Diseases & Microbiology, Europe / Italy / Verona / Negrar

Laboratory of Genomics and Bioinformatics, Comenius University Science Park, Public Health Authority of the Slovak Republic, Europe / Slovakia / Michalovce

Instituto Adolfo Lutz, Rapid Response Center, Strategic Laboratory, Instituto Adolfo Lutz, Rapid Response Center, Strategic Laboratory, South America / Brazil / Sao Paulo / Sao Paulo

Instituto Adolfo Lutz Strategic Laboratory, UBDS DR. Italo Baruffi Castelo Branco, South America / Brazil / Sao Paulo / Ribeirao Preto

Kamituga General Reference Hospital, Kamituga General Reference Hospital, Africa / Democratic Republic of the Congo / South-Kivu

Grubaugh Lab - Yale School of Public Health, Connecticut Department of Public Health, North America / USA / New York

State Research Center of Virology and Biotechnology Vector, State Research Center of Virology and Biotechnology Vector, unknown

Environmental, Agricultural, and Occupational Health, University of Nebraska Medical Center, Environmental, Agricultural, and Occupational Health, University of Nebraska Medical Center, North America / USA / Oklahoma

Highly Pathogenic Viruses, Robert Koch Institute, Centre for Biological Threats, Highly Pathogenic Viruses, Robert Koch Institute, Centre for Biological Threats, Africa / Sudan

Laboratorio de Referencia Nacional de Viruas Inmunoprevenibles. Centro Nacional de Salud Publica. Instiuto Nacional de Salud, Laboratorio de Referencia Nacional de Viruas Inmunoprevenibles. Centro Nacional de Salud Publica. Instiuto Nacional de Salud, South America / Peru / La Libertad / Trujillo / Trujillo

Instituto de Salud Publica de Chile, Genetica Molecular and Subdepartamento de Virologia ISP Chile, South America / Chile / Valparaiso / Quilpue

Instituto Adolfo Lutz Strategic Laboratory, Hosp. Municipal Dr. Jose de Carvalho Florence, South America / Brazil / Sao Paulo / Sao Jose dos Campos

Institute of Ecology and Evolution, University of Edinburgh, Nigeria Centre for Disease Control and Prevention, Africa / Nigeria / Enugu / Enugu

Department of Health, Utah Public Health Laboratory, Department of Health, Utah Public Health Laboratory, North America / USA / Utah

University of Nebraska Medical Center, Oklahoma Pathogen Genomics Consortium, Nebraska Public Health Laboratory, North America / USA / Oklahoma

Instituto Adolfo Lutz, Rapid Response Center, Strategic Laboratory, Centro de Referencia em Especialidades Central, South America / Brazil / Sao Paulo / Ribeirao Preto

Erasmus Medical Center Department of Virology, Erasmus Medical Center Department of Virology, Europe / Netherlands

Dallas County Health & Human Services Public Health Laboratory, Parkland Health and Hospital System, North America / USA / Texas / Plano

National Virus Reference Laboratory, National Virus Reference Laboratory, Europe / Ireland

Instituto Adolfo Lutz Strategic Laboratory, UBS Centro Clair Aparecida Pavan, South America / Brazil / Parana / Londrina

Instituto Adolfo Lutz, Rapid Response Center, Strategic Laboratory, Unidade de Pronto Atendimento, South America / Brazil / Sao Paulo / Vinhedo

Los Angeles County Public Health Laboratories, QUEST DIAGNOSTICS WEST HILLS, North America / USA / California

National Center for Infectious Diseases, Centers for Disease Control and Prevention, National Center for Infectious Diseases, Centers for Disease Control and Prevention, Africa / Democratic Republic of the Congo

Instituto Adolfo Lutz Strategic Laboratory, UPA Centro, South America / Brazil / Sao Paulo / Santo Andre

Laboratorio de Referencia Nacional de Viruas Inmunoprevenibles. Centro Nacional de Salud Publica. Instiuto Nacional de Salud, Laboratorio de Referencia Nacional de Viruas Inmunoprevenibles. Centro Nacional de Salud Publica. Instiuto Nacional de Salud, South America / Peru / La Libertad / Trujillo / Alto Salaverry

Tokyo Metropolitan Institute of Public Health, Tokyo Metropolitan Institute of Public Health, Asia / Japan / Tokyo

Instituto de Diagnostico y Referencia Epidemiologicos (INDRE), LESP Tabasco, North America / Mexico / Tabasco

National Institute of Health Research and Development, RSUP Dr Hasan Sadikin, Asia / Indonesia / Jawa Barat / Bandung

Institute of Ecology and Evolution, University of Edinburgh, Nigeria Centre for Disease Control and Prevention, Africa / Nigeria / Kwara

Laboratory of Virology, INMI Lazzaro Spallanzani IRCCS, Laboratory of Virology, INMI Lazzaro Spallanzani IRCCS, Europe / Italy

Instituto Adolfo Lutz Strategic Laboratory, UBS J Nordeste, South America / Brazil / Sao Paulo / Sao Paulo

Laboratorio de Referencia Nacional de Viruas Inmunoprevenibles. Centro Nacional de Salud Publica. Instiuto Nacional de Salud, Laboratorio de Referencia Nacional de Viruas Inmunoprevenibles. Centro Nacional de Salud Publica. Instiuto Nacional de Salud, South America / Peru / Lima / Lima / San Miguel

Laboratory of Respiratory Viruses and Measles, Oswaldo Cruz Institute, FIOCRUZ, Laboratorio Central de Saude Publica do Estado da Bahia (LACEN/BA), South America / Brazil / Bahia / Castro Alves

Instituto Adolfo Lutz Strategic Laboratory, Secretaria Municipal de Saude de Batatais SP, South America / Brazil / Sao Paulo / Batatais

Instituto Adolfo Lutz, Rapid Response Center, Strategic Laboratory, UBS Parque Industrial, South America / Brazil / Sao Paulo / Sao Jose do Rio Preto

Instituto Adolfo Lutz Strategic Laboratory, Hosp. Tereza de Lisieux, South America / Brazil / Bahia / Salvador

Pathogen Genomic Laboratory, Institut National de Recherche Biomedicale (INRB), Pathogen Genomic Laboratory, Institut National de Recherche Biomedicale (INRB), Africa / Democratic Republic of the Congo / Tshopo / Kisangani

Pathogen Genomic Laboratory, Institut National de Recherche Biomedicale (INRB), Pathogen Genomic Laboratory, Institut National de Recherche Biomedicale (INRB), Africa / Democratic Republic of the Congo / Sankuru / Lodja

National Center for Infectious Diseases, Centers for Disease Control and Prevention, National Center for Infectious Diseases, Centers for Disease Control and Prevention, Africa / Republic of the Congo

Department of Acute Infectious Diseases Control and Prevention, Yunnan Center for Disease Control and Prevention, Department of Acute Infectious Diseases Control and Prevention, Yunnan Center for Disease Control and Prevention, Asia / China / Yunnan

Centers for Disease Control and Prevention, Centers for Disease Control and Prevention, Africa / Gabon

INSPI-Dirección Técnica de Investigación, Desarrollo e Innovación INSPI-Centro de Referencia Nacional de Genómica, Secuenciación y Bioinformática, INSPI-Centro de Referencia Nacional de Virus Exantemáticos, Gastroentéricos y Transmitido por Vectores., South America / Ecuador / Guayas / Guayaquil

Instituto Adolfo Lutz, Rapid Response Center, Strategic Laboratory, Instituto de Infectologia Emilio Ribas, South America / Brazil / Sao Paulo / Sao Paulo

Instituto Adolfo Lutz Strategic Laboratory, Sistema de Vigilancia em Saude Viamao, South America / Brazil / Rio Grande do Sul / Viamao

Indian Council of Medical Research-National Institute of Virology, Microbial Containment Complex, Indian Council of Medical Research-National Institute of Virology, Microbial Containment Complex, Asia / India / Kerala

Laboratorio de Referencia Nacional de Viruas Inmunoprevenibles. Centro Nacional de Salud Publica. Instiuto Nacional de Salud, Laboratorio de Referencia Nacional de Viruas Inmunoprevenibles. Centro Nacional de Salud Publica. Instiuto Nacional de Salud, South America / Peru / Arequipa / Camana / Camana

Laboratório de Ecologia de Doenças Transmissíveis na Amazônia, Instituto Leônidas e Maria Deane - Fiocruz Amazônia, Laboratório Central de Saúde Pública do Amazonas - LACEN-AM, South America / Brazil / Amazonas / Manaus

Instituto Adolfo Lutz, Rapid Response Center, Strategic Laboratory, Instituto Adolfo Lutz, Rapid Response Center, Strategic Laboratory, South America / Brazil / Sao Paulo / Itanhaem

Instituto de Diagnostico y Referencia Epidemiologicos (INDRE), Laboratorio Estatal de Salud Pública Nuevo Leon, North America / Mexico / Nuevo Leon

Laboratorio Central de Salud Publica, Laboratorio Central de Salud Publica, South America / Paraguay / Central

Laboratório de Virologia Molecular - Unidade de Genômica - UFRJ, Núcleo de Enfrentamento e Estudos de Doenças Infecciosas Emergentes e Reemergentes, South America / Brazil / Rio de Janeiro

National Institute of Health, Department of Medical Sciences, Ministry of Public Health, Thailand, Vajira Hospital, Asia / Thailand / Bangkok

Instituto Adolfo Lutz Strategic Laboratory, Coordenadoria de Vigilancia em Saude - Sao Paulo, South America / Brazil / Sao Paulo / Sao Paulo

Laboratorio de Referencia Nacional de Viruas Inmunoprevenibles. Centro Nacional de Salud Publica. Instiuto Nacional de Salud, Laboratorio de Referencia Nacional de Viruas Inmunoprevenibles. Centro Nacional de Salud Publica. Instiuto Nacional de Salud, South America / Peru / Arequipa / Arequipa / Paucarpata

Laboratorio de Referencia Nacional de Viruas Inmunoprevenibles. Centro Nacional de Salud Publica. Instiuto Nacional de Salud, Laboratorio de Referencia Nacional de Viruas Inmunoprevenibles. Centro Nacional de Salud Publica. Instiuto Nacional de Salud, South America / Peru / Ica

Instituto Adolfo Lutz Strategic Laboratory, Hospital Vera Cruz, South America / Brazil / Sao Paulo / Campinas

Pathogen Genomic Laboratory, Institut National de Recherche Biomedicale, Pathogen Genomic Laboratory, Institut National de Recherche Biomedicale, Africa / Democratic Republic of the Congo / Equateur

Instituto Adolfo Lutz Strategic Laboratory, Secrtetaria Municipal de Saude de Sata Barbara D Oeste, South America / Brazil / Sao Paulo / Santa Barbara D Oeste

Instituto Adolfo Lutz Strategic Laboratory, Hosp. Sirio-Libanes, South America / Brazil / Sao Paulo / Sao Paulo

Instituto Adolfo Lutz Strategic Laboratory, UBS J COPA, South America / Brazil / Sao Paulo / Sao Paulo

Public Health Laboratory, NYC Department of Health and Mental Hygiene, Public Health Laboratory, NYC Department of Health and Mental Hygiene, North America / USA / New York / New York

Instituto Adolfo Lutz Strategic Laboratory, Unidade Basica de Saude Esplanada, South America / Brazil / Sao Paulo / Jundiai

National Institute of Health Research and Development, PKC Setiabudi, Asia / Indonesia / DKI Jakarta / Jakarta Selatan

Department of Clinical Sciences, Institute of Tropica Medicine, Department of Clinical Sciences, Institute of Tropica Medicine, Europe / Belgium

Pathogen Genomic Laboratory, Institut National de Recherche Biomedicale (INRB), Pathogen Genomic Laboratory, Institut National de Recherche Biomedicale (INRB), Africa / Democratic Republic of the Congo / Equateur

National Institute of Health Research and Development, Eka Hospital BSD, Asia / Indonesia / Banten / Tangerang Selatan

Instituto de Diagnostico y Referencia Epidemiologicos (INDRE), LESP Sinaloa, North America / Mexico / Sinaloa

National Virus Reference Laboratory, National Virus Reference Laboratory, Europe / Ireland / Donegal

Medical University of Vienna Center for Virology, Medical University of Vienna Center for Virology, Europe / Austria / Vienna

Instituto Adolfo Lutz Strategic Laboratory, Hospital Albert Sabin Atibaia, South America / Brazil / Sao Paulo / Atibaia

Guangdong Provincial Center for Disease Control and Prevention, Institute of Pathogenic Microbiology, Guangdong Provincial Center for Disease Control and Prevention, Institute of Pathogenic Microbiology, Asia / China

Pathogen Genomic Laboratory, Institut National de Recherche Biomedicale (INRB), Pathogen Genomic Laboratory, Institut National de Recherche Biomedicale (INRB), Africa / Democratic Republic of the Congo / Mai-Ndombe

Instituto Adolfo Lutz, Rapid Response Center, Strategic Laboratory, Pronto Socorro Municipal Perus, South America / Brazil / Sao Paulo / Sao Paulo

MEPHI, IHU - Mediterranee Infection, MEPHI, IHU - Mediterranee Infection, Europe / France

Laboratory Medicine and Pathology, University of Washington, Laboratory Medicine and Pathology, University of Washington, North America / USA / Washington

Laboratorio de Referencia Nacional de Viruas Inmunoprevenibles. Centro Nacional de Salud Publica. Instiuto Nacional de Salud, Laboratorio de Referencia Nacional de Viruas Inmunoprevenibles. Centro Nacional de Salud Publica. Instiuto Nacional de Salud, South America / Peru / Lima / Lima / Ancon

Hospital Universitario A Coruña, Microbiology Service, A Coruña University Hospital Complex (CHUAC), Europe / Spain / Galicia / A Coruna

Instituto Adolfo Lutz, Rapid Response Center, Strategic Laboratory, Hospital e Maternidade Santa Maria Cruz Azul, South America / Brazil / Sao Paulo / Sao Paulo

Instituto Adolfo Lutz Strategic Laboratory, Unidade Mista de saude Mariano Gayoso Castelo Branco, South America / Brazil / Piaui / Teresina

Department of immunology and microbiology - Pasteur Institute in Ho Chi Minh city, Department of immunology and microbiology - Pasteur Institute in Ho Chi Minh city, Asia / Vietnam / Ho Chi Minh City

National Institute of Health Research and Development, PKC Cakung, Asia / Indonesia / DKI Jakarta / Jakarta Timur

Instituto de Salud Publica de Chile, Genetica Molecular and Subdepartamento de Virologia ISP Chile, South America / Chile / Region Metropolitana de Santiago / Penalolen

Naval Medical Research Center Biological Defense Research Directorate, Naval Infectious Diseases Diagnostic Laboratory, North America / USA / Maryland

Laboratorio de Referencia Nacional de Virus Respiratorio. Centro Nacional de Salud Publica. Instituto Nacional de Salud., Laboratorio de Referencia Nacional de Virus Respiratorio. Centro Nacional de Salud Publica. Instituto Nacional de Salud., South America / Peru / Callao

Grubaugh Lab - Yale School of Public Health, Connecticut Department of Public Health, North America / USA / Connecticut

Instituto Adolfo Lutz Strategic Laboratory, USF Jardim Oratorio, South America / Brazil / Sao Paulo / Maua

Instituto Adolfo Lutz, Rapid Response Center, Strategic Laboratory, Centro de Referencia e Treinamento DSTAIDS, South America / Brazil / Sao Paulo / Sao Paulo

Instituto de Diagnostico y Referencia Epidemiologicos (INDRE), LESP Yucatan, North America / Mexico / Yucatan

Laboratorio de Referencia Nacional de Viruas Inmunoprevenibles. Centro Nacional de Salud Publica. Instiuto Nacional de Salud, Laboratorio de Referencia Nacional de Viruas Inmunoprevenibles. Centro Nacional de Salud Publica. Instiuto Nacional de Salud, South America / Peru / Lima / Lima / Miraflores

Instituto Adolfo Lutz, Rapid Response Center, Strategic Laboratory, Hospital Albert Einstein, South America / Brazil / Sao Paulo / Sao Paulo

Instituto Adolfo Lutz Strategic Laboratory, UMS Campina do Siqueira, South America / Brazil / Parana / Curitiba

Institute of Ecology and Evolution, University of Edinburgh, Nigeria Centre for Disease Control and Prevention, Africa / Nigeria / Rivers

Microbiology Department. Complexo Hospitalario Universitario de Vigo, Microbiology Department. Complexo Hospitalario Universitario de Vigo, Europe / Spain / Galicia / Vigo

Public and Environmental Health Reference Laboratories, Pathology Queensland, 4 Cyte Pathology, Oceania / Australia / Queensland

Pathogen Genomic Laboratory, Institut National de Recherche Biomedicale (INRB), Pathogen Genomic Laboratory, Institut National de Recherche Biomedicale (INRB), Africa / Democratic Republic of the Congo / Equateur / Gemena

Instituto Adolfo Lutz Strategic Laboratory, Vigilancia Epidemiologica Municipal, South America / Brazil / Sao Paulo / Patrocinio Paulista

Instituto Adolfo Lutz, Rapid Response Center, Strategic Laboratory, Hospital Samaritano, South America / Brazil / Sao Paulo / Sao Paulo

Bundeswehr Institute of Microbiology, Division of Infectious Diseases and Tropical Medicine, University Hospital, Ludwig-Maximilians-Universitaet (LMU) Munich, Munich, Germany, Europe / Germany / Bavaria / Munich

National Institute of Health Research and Development, PKC Pademangan, Asia / Indonesia / DKI Jakarta / Jakarta Utara

Instituto de Salud Publica de Chile, Genetica Molecular and Subdepartamento de Virologia ISP Chile, South America / Chile / Region Metropolitana de Santiago / Santiago

Instituto Adolfo Lutz Strategic Laboratory, Hosp. Municipal de Ilhabela Gov. Mario Covas Jr., South America / Brazil / Sao Paulo / Ilhabela

Instituto de Salud Publica de Chile, Genetica Molecular and Subdepartamento de Virologia ISP Chile, South America / Chile / Region Metropolitana de Santiago / Pedro Aguirre Cerda

Public and Environmental Health Reference Laboratories, Pathology Queensland, Sullivan Nicolaides Pathology, Oceania / Australia / Queensland

Research Institute for Tropical Medicine, Research Institute for Tropical Medicine, Asia / Philippines / National Capital Region

Instituto Adolfo Lutz, Rapid Response Center, Strategic Laboratory, Instituto Adolfo Lutz, Rapid Response Center, Strategic Laboratory, South America / Brazil / Sao Paulo / Jardinopolis

Laboratory of Respiratory Viruses and Measles, Oswaldo Cruz Institute, FIOCRUZ, Laboratorio de Enterovirus, Instituto Oswaldo Cruz, Fiocruz, South America / Brazil / Rio de Janeiro / Rio de Janeiro

National Institute of Health Research and Development, Dinkes Kabupaten Cirebon, Asia / Indonesia / Jawa Barat / Cirebon

Laboratorio de Referencia Nacional de Virus Inmunoprevenibles. Centro Nacional de Salud Publica. Instituto Nacional de Salud., Laboratorio de Referencia Nacional de Virus Inmunoprevenibles. Centro Nacional de Salud Publica. Instituto Nacional de Salud., South America / Peru / Lima

National Institute of Health Research and Development, PKC Kelapa Gading, Asia / Indonesia / DKI Jakarta / Jakarta Utara

Indian Council of Medical Research-National Institute of Virology, Microbial Containment Complex, Indian Council of Medical Research-National Institute of Virology, Microbial Containment Complex, Asia / India / Delhi

Instituto de Diagnostico y Referencia Epidemiologicos (INDRE), Laboratorio Estatal de Salud Pública Puebla, North America / Mexico / Puebla

Instituto Adolfo Lutz, Rapid Response Center, Strategic Laboratory, Santa Casa de Sao Paulo Hospital Santa Isabel, South America / Brazil / Sao Paulo / Sao Paulo

Laboratorio de Referencia Nacional de Viruas Inmunoprevenibles. Centro Nacional de Salud Publica. Instiuto Nacional de Salud, Laboratorio de Referencia Nacional de Viruas Inmunoprevenibles. Centro Nacional de Salud Publica. Instiuto Nacional de Salud, South America / Peru / La Libertad / Trujillo / La Esperanza

Laboratorio de Referencia Nacional de Viruas Inmunoprevenibles. Centro Nacional de Salud Publica. Instiuto Nacional de Salud, Laboratorio de Referencia Nacional de Viruas Inmunoprevenibles. Centro Nacional de Salud Publica. Instiuto Nacional de Salud, South America / Peru / Lima / Lima / La Molina

Instituto Adolfo Lutz Strategic Laboratory, Laboratorio de Vigilancia em Saude de Vinhedo, South America / Brazil / Sao Paulo / Vinhedo

National Institute of Health Research and Development, PKM Kembangan, Asia / Indonesia / DKI Jakarta / Jakarta Barat

CDCT/CEVS/SES-RS, CDCT/CEVS/SES-RS, South America / Brazil / Rio Grande do Sul / Porto Alegre

Instituto de Diagnostico y Referencia Epidemiologicos (INDRE), LESP Hidalgo, North America / Mexico / Hidalgo

Laboratorio de Referencia Nacional de Viruas Inmunoprevenibles. Centro Nacional de Salud Publica. Instiuto Nacional de Salud, Laboratorio de Referencia Nacional de Viruas Inmunoprevenibles. Centro Nacional de Salud Publica. Instiuto Nacional de Salud, South America / Peru / Callao / Callao / Callao

Los Angeles County Public Health Laboratories, Quest Diagnostics West Hills, North America / USA / California / Los Angeles County

Laboratorio de Referencia Nacional de Viruas Inmunoprevenibles. Centro Nacional de Salud Publica. Instiuto Nacional de Salud, Laboratorio de Referencia Nacional de Viruas Inmunoprevenibles. Centro Nacional de Salud Publica. Instiuto Nacional de Salud, South America / Peru / Lima / Lima / La Victoria

Centro de Referencia Nacional de Genomica, Secuenciacion y Bioinformatica GENSBIO, INSPI-CZ9, Laboratorio de Virus Exantematicos, Gastroentéricos y Otros Transmitidos por Vectores, South America / Ecuador / Pichincha

Pathogen Genomic Laboratory, Institut National de Recherche Biomedicale (INRB), Pathogen Genomic Laboratory, Institut National de Recherche Biomedicale (INRB), Africa / Democratic Republic of the Congo / Kwango

Balai Besar Laboratorium Biologi Kesehatan, RSPI Sulianti Saroso, Asia / Indonesia / DKI Jakarta / Jakarta Utara

Instituto Adolfo Lutz, Rapid Response Center, Strategic Laboratory, Instituto Adolfo Lutz, Rapid Response Center, Strategic Laboratory, South America / Brazil / Sao Paulo / Santo Andre

Institute of Ecology and Evolution, University of Edinburgh, Nigeria Centre for Disease Control and Prevention, Africa / Nigeria / Ogun

Instituto Adolfo Lutz Strategic Laboratory, CTA Centro de Testagem e Aconselhamento de Caieiras, South America / Brazil / Sao Paulo / Caieiras

Instituto Adolfo Lutz Strategic Laboratory, Centro de Saude I Albertino Affonso Jaboticabal, South America / Brazil / Sao Paulo / Jaboticabal

Molecular Epidemiology, Idaho Bureau of Laboratories, Molecular Epidemiology, Idaho Bureau of Laboratories, North America / USA / Idaho

Instituto Adolfo Lutz Strategic Laboratory, Vigilancia Epidemiologica Jardinopolis - SP, South America / Brazil / Sao Paulo / Jardinopolis

Southern Nevada Public Health Laboratory, Southern Nevada Public Health Laboratory, North America / USA / Nevada / Clark

Center of Diagnostics and Vaccine Development, Centers for Disease Control, Taiwan, Center of Diagnostics and Vaccine Development, Centers for Disease Control, Taiwan, Asia / Taiwan / Taipei

Instituto de Salud Publica de Chile, Genetica Molecular and Subdepartamento de Virologia ISP Chile, South America / Chile / Valparaiso / Viña Del Mar

Army Medical and Veterinary Research Center, Laboratory of Clinical Microbiology, Virology and Bioemergencies. ASST-Fatebenefratelli-Sacco, L.Sacco University Hospital, Europe / Italy

Submission Date, Collection date, Accession ID

Instituto Adolfo Lutz Strategic Laboratory, SAE DST AIDS Cidade Dutra, South America / Brazil / Sao Paulo / Sao Paulo

National Virus Reference Laboratory, National Virus Reference Laboratory, Europe / Ireland / Kildare

Laboratorio de Referencia Nacional de Viruas Inmunoprevenibles. Centro Nacional de Salud Publica. Instiuto Nacional de Salud, Laboratorio de Referencia Nacional de Viruas Inmunoprevenibles. Centro Nacional de Salud Publica. Instiuto Nacional de Salud, South America / Peru / La Libertad / Trujillo / Victor Larco Herrera

Bundeswehr Institute of Microbiology, Bundeswehr Institute of Microbiology, Europe / Germany

National Center for Global Health and Medicine, National Center for Global Health and Medicine, Asia / Japan / Tokyo

Institute of Ecology and Evolution, University of Edinburgh, Nigeria Centre for Disease Control and Prevention, Africa / Nigeria / Delta

Department of Virology, National Institute of Health, Pakistan Institute of Medical Sciences - PIMS, Asia / Pakistan / Islamabad

Laboratorio de Referencia Nacional de Viruas Inmunoprevenibles. Centro Nacional de Salud Publica. Instiuto Nacional de Salud, Laboratorio de Referencia Nacional de Viruas Inmunoprevenibles. Centro Nacional de Salud Publica. Instiuto Nacional de Salud, South America / Peru / Arequipa / Arequipa / Cerro Colorado

Kamituga General Reference Hospital/South-Kivu, Kamituga General Reference Hospital/South-Kivu, Africa / Democratic Republic of the Congo / South-Kivu

Institute of Ecology and Evolution, University of Edinburgh, Nigeria Centre for Disease Control and Prevention, Africa / Nigeria / Kaduna / Kaduna

Instituto Adolfo Lutz, Rapid Response Center, Strategic Laboratory, Centro de Testagem e Aconselhamento de Guarulhos, South America / Brazil / Sao Paulo / Guarulhos

Laboratorio de Referencia Nacional de Viruas Inmunoprevenibles. Centro Nacional de Salud Publica. Instiuto Nacional de Salud, Laboratorio de Referencia Nacional de Viruas Inmunoprevenibles. Centro Nacional de Salud Publica. Instiuto Nacional de Salud, South America / Peru / Lima / Lima / Magdalena del Mar

National Institute of Health Research and Development, PKC Kebayoran Baru, Asia / Indonesia / DKI Jakarta / Jakarta Selatan

Instituto Adolfo Lutz Strategic Laboratory, Hosp. Sao Joaquim - Beneficiencia Portuguesa, South America / Brazil / Sao Paulo / Sao Paulo

Laboratory of Genomics and Bioinformatics, Comenius University Science Park, Public Health Authority of the Slovak Republic, Europe / Slovakia / Levice

Balai Besar Laboratorium Biologi Kesehatan, Puskesmas Cilodong, Asia / Indonesia / Jawa Barat / Depok

Instituto de Salud Publica de Chile, Genetica Molecular and Subdepartamento de Virologia ISP Chile, South America / Chile / Region Metropolitana de Santiago

Instituto Adolfo Lutz Strategic Laboratory, Hosp. Alemao Oswaldo Cruz, South America / Brazil / Sao Paulo / Sao Paulo

Laboratorio de Referencia Nacional de Viruas Inmunoprevenibles. Centro Nacional de Salud Publica. Instiuto Nacional de Salud, Laboratorio de Referencia Nacional de Viruas Inmunoprevenibles. Centro Nacional de Salud Publica. Instiuto Nacional de Salud, South America / Peru / Lima / Lima / Lima

Instituto Adolfo Lutz, Rapid Response Center, Strategic Laboratory, Hospital e Maternidade Celso Pierro - Campinas, South America / Brazil / Sao Paulo / Campinas

Instituto Adolfo Lutz Strategic Laboratory, Secretaria Municipal de Saude de Caxias do Sul, South America / Brazil / Rio Grande do Sul / Caxias do Sul

Instituto Adolfo Lutz, Rapid Response Center, Strategic Laboratory, Secretaria de Saude Publica de Praia Grande, South America / Brazil / Sao Paulo / Praia Grande

Instituto de Diagnostico y Referencia Epidemiologicos (INDRE), LESP Veracruz, North America / Mexico / Veracruz

Center for Vectors and Infectious Diseases Research (CEVDI), National Health Institute Doutor Ricardo Jorge, IP (INSA), Center for Vectors and Infectious Diseases Research (CEVDI), National Health Institute Doutor Ricardo Jorge, IP (INSA), Europe / Portugal

National Institute of Health Research and Development, PKM Tanjung Priuk, Asia / Indonesia / DKI Jakarta / Jakarta Utara

Tokyo Metropolitan Institute of Public Health, Department of Microbiology, Tokyo Metropolitan Institute of Public Health, Department of Microbiology, Asia / Japan / Tokyo

Environment and Infectious Risks Unit, Insitut Pasteur, Environment and Infectious Risks Unit, Insitut Pasteur, Europe / France

Charité Universitätsmedizin Berlin, Institute for Virology, Charité Universitätsmedizin Berlin, Institute for Virology/Laboratory Berlin, Europe / Germany / Berlin

Vaccine and Gene Therapy Institute, Oregon Health and Science University, Vaccine and Gene Therapy Institute, Oregon Health and Science University, Africa / Democratic Republic of the Congo

Laboratorio de Referencia Nacional de Viruas Inmunoprevenibles. Centro Nacional de Salud Publica. Instiuto Nacional de Salud, Laboratorio de Referencia Nacional de Viruas Inmunoprevenibles. Centro Nacional de Salud Publica. Instiuto Nacional de Salud, South America / Peru / Callao / Callao / Bellavista

Instituto Adolfo Lutz Strategic Laboratory, Casa de Saude Stella Maris, South America / Brazil / Sao Paulo / Caraguatatuba

Laboratorio de Referencia Nacional de Viruas Inmunoprevenibles. Centro Nacional de Salud Publica. Instiuto Nacional de Salud, Laboratorio de Referencia Nacional de Viruas Inmunoprevenibles. Centro Nacional de Salud Publica. Instiuto Nacional de Salud, South America / Peru / Lima / Lima / San Juan de Lurigancho

Instituto Adolfo Lutz Strategic Laboratory, Unidade Basica de Saude Vila Cristina, South America / Brazil / Sao Paulo / Piracicaba

Department of Virology, National Institute of Health, Islamabad, Pakistan, Department of Virology, National Institute of Health, Islamabad, Pakistan, Asia / Pakistan / Islamabad

Balai Besar Laboratorium Biologi Kesehatan, PKC Tebet, Asia / Indonesia / DKI Jakarta / Jakarta Selatan

Instituto Adolfo Lutz, Rapid Response Center, Strategic Laboratory, Santa Casa de Vinhedo, South America / Brazil / Sao Paulo / Vinhedo

Instituto Adolfo Lutz, Rapid Response Center, Strategic Laboratory, CR DST Aids Santo Amaro, South America / Brazil / Sao Paulo / Sao Paulo

Instituto Adolfo Lutz, Rapid Response Center, Strategic Laboratory, Instituto Adolfo Lutz, Rapid Response Center, Strategic Laboratory, South America / Brazil / Sao Paulo / Osasco

Research and Evaluation, UKHSA, Research and Evaluation, UKHSA, Europe / United Kingdom

Laboratory of Virology, National Institute of Allergy and Infectious Diseases (NIAID), National Institutes of Health (NIH), Laboratoire National de Santé Publique (LNSP), Africa / Republic of the Congo / Likouala

National Institute of Health Research and Development, PKC Cengkareng, Asia / Indonesia / DKI Jakarta / Jakarta Barat

Public Health Laboratory, NYC Department of Health and Mental Hygiene, Public Health Laboratory, NYC Department of Health and Mental Hygiene, North America / USA / Ney York / New York City

Queensland Medical Laboratories, Oceania / Australia / Queensland, 197,195

Instituto Adolfo Lutz Strategic Laboratory, Servico de Vigilancia Epidemiologica e de Zoonoses do Guaruja, South America / Brazil / Sao Paulo / Guaruja

Korea Disease Control and Prevention Agency, Korea Disease Control and Prevention Agency, Asia / South Korea

Instituto de Diagnostico y Referencia Epidemiologicos (INDRE), LESP Quintana Roo, North America / Mexico / Quintana Roo

Laboratorio de Referencia Nacional de Virus Inmunoprevenibles. Centro Nacional de Salud Publica. Instituto Nacional de Salud., Laboratorio de Referencia Nacional de Virus Inmunoprevenibles. Centro Nacional de Salud Publica. Instituto Nacional de Salud., South America / Peru / La Libertad

Centro de Investigacion e Innovacion en Virologia Medica, Departamento de Bioquimica y Medicina Molecular, Facultad de Medicina, Universidad Autonoma de Nuevo Leon, Servicio de Infectologia, Hospital Universitario Dr. José Eleuterio Gonzalez, Universidad Autonoma de Nuevo Leon, North America / Mexico / Nuevo Leon

Pathogen Genomics Lab, National Institute for Biomedical Research (INRB), Pathogen Genomics Lab, National Institute for Biomedical Research (INRB), Africa / Democratic Republic of the Congo

Instituto de Salud Publica de Chile, Genetica Molecular and Subdepartamento de Virologia ISP Chile, South America / Chile / Region Metropolitana de Santiago / La Pintana

Direccion de Investigacion en Salud Publica, Instituto Nacional de Salud, Direccion de Investigacion en Salud Publica, Instituto Nacional de Salud, South America / Colombia / Cundinamarca

Instituto Adolfo Lutz Strategic Laboratory, Centro de Saude Gabriel de Lara, South America / Brazil / Parana / Paranagua

Instituto Adolfo Lutz Strategic Laboratory, PSF Vila Nossa Senhora de Fatima Fartura, South America / Brazil / Sao Paulo / Fartura

Instituto Adolfo Lutz, Rapid Response Center, Strategic Laboratory, Unidade Basica de Saude Luciano Rodrigues Costa, South America / Brazil / Sao Paulo / Osasco

National Virus Reference Laboratory, St Jame's Hospital, Virology Department, Europe / Ireland / Dublin

Instituto de Salud Publica de Chile, Genetica Molecular and Subdepartamento de Virologia ISP Chile, South America / Chile / Region Metropolitana de Santiago / Providencia

Instituto Adolfo Lutz, Rapid Response Center, Strategic Laboratory, Servico de Atendimento Especializado Saecrt Hivaids, South America / Brazil / Sao Paulo / Sao Jose do Rio Preto

National Virus Reference Laboratory, National Virus Reference Laboratory, Europe / Ireland / Laois

Research and Evaluation, United Kingdom Health Security Agency, Research and Evaluation, United Kingdom Health Security Agency, Europe / United Kingdom

Instituto Adolfo Lutz Strategic Laboratory, Hosp. Itacolomy Butanta, South America / Brazil / Sao Paulo / Itapevi

Laboratorio de Referencia Nacional de Viruas Inmunoprevenibles. Centro Nacional de Salud Publica. Instiuto Nacional de Salud, Laboratorio de Referencia Nacional de Viruas Inmunoprevenibles. Centro Nacional de Salud Publica. Instiuto Nacional de Salud, South America / Peru / Piura / Piura / Piura

Laboratorio de Referencia Nacional de Viruas Inmunoprevenibles. Centro Nacional de Salud Publica. Instiuto Nacional de Salud, Laboratorio de Referencia Nacional de Viruas Inmunoprevenibles. Centro Nacional de Salud Publica. Instiuto Nacional de Salud, South America / Peru / Lima / Lima / Carabayllo

Environmental, Agricultural, and Occupational Health, University of Nebraska Medical Center, Environmental, Agricultural, and Occupational Health, University of Nebraska Medical Center, North America / USA / Nebraska

National Institute for Infectious Diseases 'Lazzaro Spallanzani', Laboratory of Virology, National Institute for Infectious Diseases 'Lazzaro Spallanzani', Laboratory of Virology, Europe / Italy

National Institute of Health Research and Development, PKC Tanah Abang, Asia / Indonesia / DKI Jakarta / Jakarta Pusat

National Institute of Health Research and Development, RSUPN Dr Cipto Mangunkusumo, Asia / Indonesia / DKI Jakarta / Jakarta Pusat

Virology, GENomique EPIdemiologique des maladies Infectieuses, Virology, GENomique EPIdemiologique des maladies Infectieuses, Europe / France

Instituto Adolfo Lutz, Rapid Response Center, Strategic Laboratory, Unidade Basica de Saude Maria Girade Cury, South America / Brazil / Sao Paulo / Osasco

Hospital Universitario La Paz, Microbiology, Hospital Universitario La Paz, Microbiology, Europe / Spain / Madrid

Biochemistry and Molecular Biology, Israel Institute for Biological Research, Biochemistry and Molecular Biology, Israel Institute for Biological Research, Asia / Israel

Institut Pasteur, Bichat-Claude Bernard Hospital, Paris France, Europe / France

Laboratorio de Referencia Nacional de Viruas Inmunoprevenibles. Centro Nacional de Salud Publica. Instiuto Nacional de Salud, Laboratorio de Referencia Nacional de Viruas Inmunoprevenibles. Centro Nacional de Salud Publica. Instiuto Nacional de Salud, South America / Peru / Arequipa / Arequipa / Cayma

Instituto de Salud Publica de Chile, Genetica Molecular and Subdepartamento de Virologia ISP Chile, South America / Chile / Arica y Parinacota / Arica

Institute of Ecology and Evolution, University of Edinburgh, Nigeria Centre for Disease Control and Prevention, Africa / Nigeria / Bayelsa

Instituto de Salud Publica de Chile, Genetica Molecular and Subdepartamento de Virologia ISP Chile, South America / Chile / Region Metropolitana de Santiago / San Bernardo

Instituto de Diagnostico y Referencia Epidemiologicos (INDRE), LESP Mexico City, North America / Mexico / Mexico City

Research and Evaluation, UKHSA, Research and Evaluation, UKHSA, Europe / United Kingdom / England

Instituto Adolfo Lutz Strategic Laboratory, Santa Casa de Atibaia Pro Saude, South America / Brazil / Sao Paulo / Atibaia

Institute of Ecology and Evolution, University of Edinburgh, Nigeria Centre for Disease Control and Prevention, Africa / Nigeria / Imo

Laboratorio de Referencia Nacional de Viruas Inmunoprevenibles. Centro Nacional de Salud Publica. Instiuto Nacional de Salud, Laboratorio de Referencia Nacional de Viruas Inmunoprevenibles. Centro Nacional de Salud Publica. Instiuto Nacional de Salud, South America / Peru / Callao / Callao / Ventanilla

Chemical, Biological and Radiological Sciences, Defence Science and Technology Laboratory, Nigeria Centre for Disease Control, National Reference Laboratory, Africa / Nigeria

Laboratory of Respiratory Viruses and Measles, Oswaldo Cruz Institute, FIOCRUZ, Laboratorio Central de Saude Publica do Estado da Bahia (LACEN/BA), South America / Brazil / Bahia / Maracas

Centers for Disease Control and Prevention, Centers for Disease Control and Prevention, Africa / Nigeria

Los Angeles County Public Health Laboratories, Los Angeles County Public Health Laboratory, North America / USA / California / Los Angeles County

Instituto de Diagnostico y Referencia Epidemiologicos (INDRE), LESP Aguascalientes, North America / Mexico / Aguascalientes

Medical Microbiology & Infection Prevention, Amsterdam Medical Centres location AMC, Medical Microbiology & Infection Prevention, Amsterdam Medical Centres location AMC, Europe / Netherlands / Noord-Holland / Amsterdam

Laboratorio de Referencia Nacional de Viruas Inmunoprevenibles. Centro Nacional de Salud Publica. Instiuto Nacional de Salud, Laboratorio de Referencia Nacional de Viruas Inmunoprevenibles. Centro Nacional de Salud Publica. Instiuto Nacional de Salud, South America / Peru / Lima / Lima / Surquillo

Instituto Adolfo Lutz Strategic Laboratory, CRT-DST-AIDS, South America / Brazil / Sao Paulo / Sao Paulo

Instituto de Salud Publica de Chile, Genetica Molecular and Subdepartamento de Virologia ISP Chile, South America / Chile / Region Metropolitana de Santiago / Lo Espejo

National Institute of Health Research and Development, PKM Warung Jambu, Asia / Indonesia / Jawa Barat / Bogor

Instituto Adolfo Lutz Strategic Laboratory, Secretaria Municipal de Saude de Suzano, South America / Brazil / Sao Paulo / Suzano

Instituto de Diagnostico y Referencia Epidemiologicos (INDRE), LESP Tlaxcala, North America / Mexico / Tlaxcala

Instituto de Diagnostico y Referencia Epidemiologicos (INDRE), LESP Guanajuato, North America / Mexico / Guanajuato

Laboratory of Respiratory Viruses and Measles, Oswaldo Cruz Institute, FIOCRUZ, Laboratorio Central de Saude Publica do Estado da Bahia (LACEN/BA), South America / Brazil / Bahia / Conceicao da Feira

Instituto de Salud Publica de Chile, Genetica Molecular and Subdepartamento de Virologia ISP Chile, South America / Chile / Region Metropolitana de Santiago / Ñuñoa

Virology, APHP Pitie Salpetriere SU, Virology, APHP Pitie Salpetriere SU, Europe / France

Laboratoire National de Reference, Institut National de Sante Publique of Burundi, Laboratoire National de Reference, Institut National de Sante Publique of Burundi, Africa / Burundi / Kayanza

Uganda Virus Research Institute, MRC/UVRI & LSHTM Uganda Research Unit, Uganda Virus Research Institute, MRC/UVRI & LSHTM Uganda Research Unit, Africa / Uganda / Central Region

Instituto Adolfo Lutz Strategic Laboratory, CTA Sao Miguel, South America / Brazil / Sao Paulo / Sao Paulo

National Institute of Health Research and Development, Puskesmas Bambu Apus, Asia / Indonesia / Banten / Tangerang Selatan

Instituto Oswaldo Cruz FIOCRUZ - Laboratory of Respiratory Viruses and Measles (LVRS), Laboratorio de Enterovirus, Instituto Oswaldo Cruz, Fiocruz, South America / Brazil / Rio de Janeiro / Marica

Instituto Adolfo Lutz, Rapid Response Center, Strategic Laboratory, Vigilancia Epidemiologica de Sao Bernardo do Campo, South America / Brazil / Sao Paulo / Sao Bernardo do Campo

Laboratorio de Referencia Nacional de Viruas Inmunoprevenibles. Centro Nacional de Salud Publica. Instiuto Nacional de Salud, Laboratorio de Referencia Nacional de Viruas Inmunoprevenibles. Centro Nacional de Salud Publica. Instiuto Nacional de Salud, South America / Peru / Ica / Nazca / Marcona

Institute of Microbiology, Universidad San Francisco de Quito, Institute of Microbiology, Universidad San Francisco de Quito, South America / Ecuador / Pichincha / Quito

Instituto Adolfo Lutz Strategic Laboratory, AMA Paraisopolis, South America / Brazil / Sao Paulo / Sao Paulo

National Virus Reference Laboratory, National Virus Reference Laboratory, Europe / Ireland / Limerick

Laboratorio de Referencia Nacional de Viruas Inmunoprevenibles. Centro Nacional de Salud Publica. Instiuto Nacional de Salud, Laboratorio de Referencia Nacional de Viruas Inmunoprevenibles. Centro Nacional de Salud Publica. Instiuto Nacional de Salud, South America / Peru / Lima / Lima / Villa Maria del Triunfo

Instituto Adolfo Lutz, Rapid Response Center, Strategic Laboratory, Instituto Adolfo Lutz, Rapid Response Center, Strategic Laboratory, South America / Brazil / Sao Paulo / Sao Jose Dos Campos

Instituto Adolfo Lutz Strategic Laboratory, Unidade Mista de Atendimento Infantil Carapicuiba, South America / Brazil / Sao Paulo / Carapicuiba

Instituto Adolfo Lutz, Rapid Response Center, Strategic Laboratory, SAE DST AIDS Cidade Dutra, South America / Brazil / Sao Paulo / Sao Paulo

Laboratory of Respiratory Viruses and Measles, Oswaldo Cruz Institute, FIOCRUZ, Laboratory of Respiratory Viruses, Exanthematous, Enteroviruses and Viral Emergencies, South America / Brazil / Rio de Janeiro / Rio de Janeiro

Instituto de Diagnostico y Referencia Epidemiologicos (INDRE), LESP Nuevo Leon, North America / Mexico / Nuevo Leon

Medical University of Vienna Center for Virology, Center for Virology, Medical University of Vienna, Europe / Austria / Vienna

Laboratory of Microbiology and Virology, Ospedale Amedeo di Savoia, ASL "Città di Torino", Laboratory of Microbiology and Virology, Ospedale Amedeo di Savoia, ASL "Città di Torino", Europe / Italy

Medical University of Vienna Center for Virology, Medical Center of Vienna Center for Virology, Europe / Austria / Graz

Virology Section, Division of Microbiology,Osaka Institute of Public Health, Virology Section, Division of Microbiology,Osaka Institute of Public Health, Asia / Japan / Osaka

Instituto Adolfo Lutz, Rapid Response Center, Strategic Laboratory, Ambulatorio de Ref de Molestias Infecto Contagiosas, South America / Brazil / Sao Paulo / Santo Andre

Institute of Ecology and Evolution, University of Edinburgh, Nigeria Centre for Disease Control and Prevention, Africa / Nigeria / Akwa Ibom

Laboratorio de Referencia Nacional de Viruas Inmunoprevenibles. Centro Nacional de Salud Publica. Instiuto Nacional de Salud, Laboratorio de Referencia Nacional de Viruas Inmunoprevenibles. Centro Nacional de Salud Publica. Instiuto Nacional de Salud, South America / Peru / Lima / Lima / Lurin

Instituto de Diagnostico y Referencia Epidemiologicos (INDRE), LESP Morelos, North America / Mexico / Morelos

Institute for Virology, Philipps-University Marburg, Institute for Virology, Philipps-University Marburg, Europe / Germany / Hessen

Microbiology, Immunology and Transplantation, KU Leuven, Microbiology, Immunology and Transplantation, KU Leuven, Europe / Belgium

National Medical Center, National Medical Center, Asia / South Korea / Seoul

Instituto Adolfo Lutz, Rapid Response Center, Strategic Laboratory, PS e Maternidade Nair Fonseca Leitao Arantes, South America / Brazil / Sao Paulo / Barueri

Laboratorio de Referencia Nacional de Viruas Inmunoprevenibles. Centro Nacional de Salud Publica. Instiuto Nacional de Salud, Laboratorio de Referencia Nacional de Viruas Inmunoprevenibles. Centro Nacional de Salud Publica. Instiuto Nacional de Salud, South America / Peru / Loreto / Maynas / San Juan Bautista

Chongqing Municipal Center for Disease Control and Prevention, Chongqing Municipal Center for Disease Control and Prevention, Asia / China / Chongqing Municipality

Instituto Adolfo Lutz, Rapid Response Center, Strategic Laboratory, Centro de Referencia em Especialidades Central Rib Preto, South America / Brazil / Sao Paulo / Ribeirao Preto

Thai Red Cross Emerging Infectious Diseases Clinical Center and Faculty of Medicine, Chulalongkorn University, Department of Disease Control, Ministry of Public Health, Asia / Thailand

Centro de Desenvolvimento Científico e Tecnológico (CDCT), Centro Estadual de Vigilância em Saúde (CEVS) da Secretaria Estadual da Saúde (SES-RS), Centro de Desenvolvimento Científico e Tecnológico (CDCT), Centro Estadual de Vigilância em Saúde (CEVS) da Secretaria Estadual da Saúde (SES-RS), South America / Brazil / Rio Grande do Sul / Sao Leopoldo

Instituto Adolfo Lutz Strategic Laboratory, Vigilancia Epidemiologica e Controle de Vetores de Pirassununga, South America / Brazil / Sao Paulo / Pirassununga

Instituto Adolfo Lutz Strategic Laboratory, NotreDame Intermedica Saude, South America / Brazil / Sao Paulo / Osasco

Instituto Adolfo Lutz, Rapid Response Center, Strategic Laboratory, UPA Vergueiro, South America / Brazil / Sao Paulo / Sao Paulo

Laboratorio de Referencia Nacional de Viruas Inmunoprevenibles. Centro Nacional de Salud Publica. Instiuto Nacional de Salud, Laboratorio de Referencia Nacional de Viruas Inmunoprevenibles. Centro Nacional de Salud Publica. Instiuto Nacional de Salud, South America / Peru / Madre de Dios / Tambopata / Tambopata

Naval Medical Research Center Biological Defense Research Directorate, Naval Infectious Diseases Diagnostic Laboratory, North America / USA / Virginia

Instituto de Salud Publica de Chile, Genetica Molecular and Subdepartamento de Virologia ISP Chile, South America / Chile / Antofagasta / Antofagasta

Institute of Ecology and Evolution, University of Edinburgh, Nigeria Centre for Disease Control and Prevention, Africa / Nigeria / Nasarawa / Nasarawa

Spiez Laboratory, Institut National de Recherche Biomédical, Africa / Democratic Republic of the Congo

Laboratorio de Referencia Nacional de Virus Inmunoprevenibles. Centro Nacional de Salud Publica. Instituto Nacional de Salud., Laboratorio de Referencia Nacional de Virus Inmunoprevenibles. Centro Nacional de Salud Publica. Instituto Nacional de Salud., South America / Peru / Piura

National Virus Reference Laboratory, National Virus Reference Laboratory, Europe / Ireland / Waterford

National Institute of Health Research and Development, PKM Mampang Prapatan, Asia / Indonesia / DKI Jakarta / Jakarta Selatan

Department for Virology, Molecular Biology and Genome Research, R. G. Lugar Center for Public Health Research, National Center for Disease Control and Public Health (NCDC) of Georgia, Department for Virology, Molecular Biology and Genome Research, R. G. Lugar Center for Public Health Research, National Center for Disease Control and Public Health (NCDC) of Georgia, Asia / Georgia / Tbilisi

National Virus Reference Laboratory, National Virus Reference Laboratory, Europe / Ireland / Dublin

Laboratory of Respiratory Viruses and Measles, Oswaldo Cruz Institute, FIOCRUZ, Laboratory of Respiratory Viruses, Exanthematous, Enteroviruses and Viral Emergencies, South America / Brazil / Santa Catarina / Balneario Camboriu

Instituto Adolfo Lutz Strategic Laboratory, CEDIC CTA, South America / Brazil / Sao Paulo / Valinhos

Molecular Biology, Microbiology, and Biochemistry, Southern Illinois University, Molecular Biology, Microbiology, and Biochemistry, Southern Illinois University, North America / USA / Illinois

National Institute for Viral Disease Control and Prevention (IVDC), Chinese Center for Disease Control and Prevention , Beijing, China, National Institute for Viral Disease Control and Prevention (IVDC), Chinese Center for Disease Control and Prevention , Beijing, China, Asia / China / Chongqing

Instituto de Diagnostico y Referencia Epidemiologicos (INDRE), LESP Campeche, North America / Mexico / Campeche

Hôpital Général de Référence de Kamituga, Hôpital Général de Référence de Kamituga, Africa / Democratic Republic of the Congo / South Kivu

University of Victoria, Biochemistry and Microbiology, University of Victoria, Biochemistry and Microbiology, Africa / Democratic Republic of the Congo

Instituto de Diagnostico y Referencia Epidemiologicos (INDRE), LESP Tamaulipas, North America / Mexico / Tamaulipas

Instituto Adolfo Lutz, Rapid Response Center, Strategic Laboratory, Centro de Referencia e Treinamento Dstaids Sao Paulo, South America / Brazil / Sao Paulo / Sao Paulo

Pathogen Genomic Laboratory, Institut National de Recherche Biomedicale (INRB), Pathogen Genomic Laboratory, Institut National de Recherche Biomedicale (INRB), Africa / Democratic Republic of the Congo / Kwilu

Institute of Ecology and Evolution, University of Edinburgh, Nigeria Centre for Disease Control and Prevention, Africa / Nigeria / Edo

Department of Genetics, University of North Carolina at Chapel Hill, Department of Genetics, University of North Carolina at Chapel Hill, North America / USA / North Carolina

Balai Besar Laboratorium Biologi Kesehatan, PKC Menteng, Asia / Indonesia / DKI Jakarta / Jakarta Pusat

Instituto de Salud Publica de Chile, Genetica Molecular and Subdepartamento de Virologia ISP Chile, South America / Chile / Region Metropolitana de Santiago / Estacion Central

RIPHL at Rush University Medical Center, Rush University Medical Center, North America / USA / Illinois / Cook County / Chicago

Department of Infectious Diseases, National Institute of Health Doutor Ricardo Jorge (INSA), Department of Infectious Diseases, National Institute of Health Doutor Ricardo Jorge (INSA), Europe / Portugal

Thai Red Cross Emerging Infectious Diseases Clinical Center and Faculty of Medicine, Chulalongkorn University, Phuket Provincial Public Health Office, Asia / Thailand

Instituto Adolfo Lutz Strategic Laboratory, Pronto Atendimento Infantil e entral de Quimioterapia SJrpreto, South America / Brazil / Sao Paulo / Sao Jose do Rio Preto

Unidade de Genômica - UFRJ, Unidade de Genômica - UFRJ, South America / Brazil / Rio de Janeiro

Oregon Health and Science University, Vaccine and Gene Therapy Institute, Oregon Health and Science University, Vaccine and Gene Therapy Institute, unknown

Instituto Adolfo Lutz Strategic Laboratory, Pronto Socorro Central de Diadema, South America / Brazil / Sao Paulo / Diadema

Instituto Adolfo Lutz Strategic Laboratory, Santa Casa de Barretos, South America / Brazil / Sao Paulo / Barretos

National Institute of Health Research and Development, RS Grha Kedoya Jakarta, Asia / Indonesia / DKI Jakarta / Jakarta Barat

Laboratório de Virologia Molecular - Unidade de Genômica - UFRJ, Laboratóro Central Rio de Janeiro, South America / Brazil / Rio de Janeiro

Dallas County Health & Human Services Public Health Laboratory, Parkland Health and Hospital System, North America / USA / Texas / Dallas

Laboratorio Nacional de Salud Pública Dr. Defilló, Laboratorio Nacional de Salud Pública Dr. Defilló, North America / Dominican Republic / Santo Domingo

Instituto Adolfo Lutz Strategic Laboratory, NotreDame Intermedica Saude Santo Andre, South America / Brazil / Sao Paulo / Santo Andre

Laboratorio Central, Ministerio de Salud Córdoba, Laboratorio Central, Ministerio de Salud Córdoba, South America / Argentina / Cordoba / Cordoba

Laboratorio de Referencia Nacional de Viruas Inmunoprevenibles. Centro Nacional de Salud Publica. Instiuto Nacional de Salud, Laboratorio de Referencia Nacional de Viruas Inmunoprevenibles. Centro Nacional de Salud Publica. Instiuto Nacional de Salud, South America / Peru / Ucayali / Coronel Portillo / Campoverde

National Virus Reference Laboratory, National Virus Reference Laboratory, Europe / Ireland / Galway

The Peter Doherty Institute for Infection and Immunity at the University of Melbourne, Department of Infectious Diseases, The Peter Doherty Institute for Infection and Immunity at the University of Melbourne, Department of Infectious Diseases, Oceania / Australia / Victoria

Instituto Adolfo Lutz Strategic Laboratory, Laboratório Central de Saúde Pública do Estado do Rio Grande do Sul, South America / Brazil / Rio Grande do Sul / Porto Alegre

Instituto Adolfo Lutz Strategic Laboratory, Secretaria Minicipal de Saude de IRECE, South America / Brazil / Bahia / Irece

Instituto Adolfo Lutz Strategic Laboratory, Hosp. Municipal Dr. Waldemar Tebaldi, South America / Brazil / Sao Paulo / Americana

State Research Center of Virology and Biotechnology Vector, State Research Center of Virology and Biotechnology Vector, Africa / Zaire / Sankuru / Kasai Oriental

Institute of Ecology and Evolution, University of Edinburgh, Nigeria Centre for Disease Control and Prevention, Africa / Nigeria / Lagos / Lagos

Laboratory of Genomics and Bioinformatics, Comenius University Science Park, Public Health Authority of the Slovak Republic, Europe / Slovakia / Dunajska Streda

Instituto Adolfo Lutz Strategic Laboratory, Hosp. Municipal Enf. Antonio Policarpo de Oliveira, South America / Brazil / Sao Paulo / Cajamar

Instituto de Diagnostico y Referencia Epidemiologicos (INDRE), LESP State of Mexico, North America / Mexico / State of Mexico

Los Angeles County Public Health Laboratories, Cedars-Sinai Medical Center, North America / USA / California / Los Angeles County

Los Angeles County Public Health Laboratories, Los Angeles County Public Health Laboratories, North America / USA / California / Los Angeles County

Fumi Kasuya Tokyo Metropolitan Institute of Public Health, Department of Microbiology, Fumi Kasuya Tokyo Metropolitan Institute of Public Health, Department of Microbiology, Asia / Japan / Tokyo

Laboratorio de Referencia Nacional de Viruas Inmunoprevenibles. Centro Nacional de Salud Publica. Instiuto Nacional de Salud, Laboratorio de Referencia Nacional de Viruas Inmunoprevenibles. Centro Nacional de Salud Publica. Instiuto Nacional de Salud, South America / Peru / Lima / Lima / Santiago de Surco

Medical University of Vienna Center for Virology, Center for Virology, Medical University of Vienna, Europe / Austria / Lower Austria

Pathogen Genomic Laboratory, Institut National de Recherche Biomedicale, Pathogen Genomic Laboratory, Institut National de Recherche Biomedicale, Africa / Democratic Republic of the Congo / Tshopo / Basoko

National Institute of Health Research and Development, PKC Mampang Prapatan, Asia / Indonesia / DKI Jakarta / Jakarta Selatan

Laboratorio Central de Salud Publica, Laboratorio Central de Salud Publica, South America / Paraguay / Capital

Public and Environmental Health Reference Laboratories, Pathology Queensland, Pathology Queensland, Oceania / Australia / Queensland

Pathogen Genomic Laboratory, Institut National de Recherche Biomedicale (INRB), Pathogen Genomic Laboratory, Institut National de Recherche Biomedicale (INRB), Africa / Democratic Republic of the Congo / Haut-UÃ©lÃ© / Isiro

Pathogen Genomic Laboratory, Institut National de Recherche Biomedicale, Pathogen Genomic Laboratory, Institut National de Recherche Biomedicale, Africa / Democratic Republic of the Congo / Kinshasa / Kinshasa

Laboratorio de Referencia Nacional de Viruas Inmunoprevenibles. Centro Nacional de Salud Publica. Instiuto Nacional de Salud, Laboratorio de Referencia Nacional de Viruas Inmunoprevenibles. Centro Nacional de Salud Publica. Instiuto Nacional de Salud, South America / Peru / San Martin / Lamas / Zapatero

Pathogen Genomic Laboratory, Institut National de Recherche Biomedicale (INRB), Pathogen Genomic Laboratory, Institut National de Recherche Biomedicale (INRB), Africa / Democratic Republic of the Congo / Tshopo / Basoko

Laboratorio de Referencia Nacional de Viruas Inmunoprevenibles. Centro Nacional de Salud Publica. Instiuto Nacional de Salud, Laboratorio de Referencia Nacional de Viruas Inmunoprevenibles. Centro Nacional de Salud Publica. Instiuto Nacional de Salud, South America / Peru / Lima / Lima / Comas

Laboratorio de Referencia Nacional de Viruas Inmunoprevenibles. Centro Nacional de Salud Publica. Instiuto Nacional de Salud, Laboratorio de Referencia Nacional de Viruas Inmunoprevenibles. Centro Nacional de Salud Publica. Instiuto Nacional de Salud, South America / Peru / Piura / Piura / 26 de Octubre

National Institute for Communicable Diseases of the National Health Laboratory Service, National Institute for Communicable Diseases of the National Health Laboratory Service, Africa / South Africa / Gauteng

Instituto de Salud Publica de Chile, Genetica Molecular and Subdepartamento de Virologia ISP Chile, South America / Chile / Valparaiso / Nogales

Instituto Adolfo Lutz, Rapid Response Center, Strategic Laboratory, Hospital de Clinicas Sul - Sao Jose dos Campos, South America / Brazil / Sao Paulo / Sao Jose Dos Campos

Instituto de Salud Publica de Chile, Genetica Molecular and Subdepartamento de Virologia ISP Chile, South America / Chile / Region Metropolitana de Santiago / Huechuraba

Indian Council of Medical Research-National Institute of Virology, Indian Council of Medical Research-National Institute of Virology, Asia / India / Kerala

Thai Red Cross Emerging Infectious Diseases Clinical Center and Faculty of Medicine, Chulalongkorn University, Bangkok Hospital Phuket, Asia / Thailand

Instituto Adolfo Lutz Strategic Laboratory, SAE DST / Aids Ipiranga Jose Francisco Araujo, South America / Brazil / Sao Paulo / Sao Paulo

National Institute of Health Research and Development, RSUD Kembangan, Asia / Indonesia / DKI Jakarta / Jakarta Barat

Pathogen Genomic Laboratory, Institut National de Recherche Biomedicale (INRB), Pathogen Genomic Laboratory, Institut National de Recherche Biomedicale (INRB), Africa / Democratic Republic of the Congo / Kinshasa

Instituto Adolfo Lutz, Rapid Response Center, Strategic Laboratory, Secretaria Municipal de Saude de Suzano, South America / Brazil / Sao Paulo / Suzano

Instituto Adolfo Lutz, Rapid Response Center, Strategic Laboratory, Secretaria Municipal de Saude Sorocaba, South America / Brazil / Sao Paulo / Sorocaba

Hangzhou Center for Disease Control and Prevention, Hangzhou Center for Disease Control and Prevention, Asia / China / Zhejiang / Hangzhou

National Institute of Health Research and Development, PKC Jatinegara, Asia / Indonesia / DKI Jakarta / Jakarta Timur

Instituto Adolfo Lutz, Rapid Response Center, Strategic Laboratory, Instituto Adolfo Lutz, Rapid Response Center, Strategic Laboratory, South America / Brazil / Sao Paulo / Praia Grande

RIPHL at Rush University Medical Center, Northwestern Medicine, North America / USA / Illinois / Cook County / Chicago

Laboratorio de Referencia Nacional de Viruas Inmunoprevenibles. Centro Nacional de Salud Publica. Instiuto Nacional de Salud, Laboratorio de Referencia Nacional de Viruas Inmunoprevenibles. Centro Nacional de Salud Publica. Instiuto Nacional de Salud, South America / Peru / Tacna / Tacna / Coronel Gregorio Albarracin

Instituto Adolfo Lutz, Rapid Response Center, Strategic Laboratory, Hospital Sao Paulo, South America / Brazil / Sao Paulo / Sao Paulo

Laboratorio de Referencia Nacional de Virus Respiratorio. Centro Nacional de Salud Publica. Instituto Nacional de Salud., Laboratorio de Referencia Nacional de Virus Respiratorio. Centro Nacional de Salud Publica. Instituto Nacional de Salud., South America / Peru / Cusco

Instituto Adolfo Lutz Strategic Laboratory, Laboratorio Municipal de Piracicaba, South America / Brazil / Sao Paulo / Piracicaba

Laboratory of Respiratory Viruses and Measles, Oswaldo Cruz Institute, FIOCRUZ, Laboratory of Respiratory Viruses, Exanthematous, Enteroviruses and Viral Emergencies, South America / Brazil / Santa Catarina / Florianopolis

National Institute of Health Research and Development, PKM Bogor Timur, Asia / Indonesia / Jawa Barat / Bogor

Instituto de Diagnóstico y Referencia Epidemiológicos/Instituto de Biotecnología UNAM, Instituto de Diagnóstico y Referencia Epidemiológicos/Jurisdicción Sanitaria Cuauhtémoc/Hospital Ángeles Roma, North America / Mexico / Mexico City

Molecular Biology Laboratory, Research Institute for Tropical Medicine, Eastwood Medical City, Asia / Philippines / National Capital Region

Instituto de Diagnostico y Referencia Epidemiologicos (INDRE), LESP Puebla, North America / Mexico / Puebla

Instituto Adolfo Lutz, Rapid Response Center, Strategic Laboratory, CR DST AIDS Santo Amaro, South America / Brazil / Sao Paulo / Sao Paulo

Instituto de Salud Publica de Chile, Genetica Molecular and Subdepartamento de Virologia ISP Chile, South America / Chile / O'Higgins / Rengo

United States Army Medical Research Institute of Infectious Diseases, Center for Genome Sciences, United States Army Medical Research Institute of Infectious Diseases, Center for Genome Sciences, unknown

Instituto Adolfo Lutz, Rapid Response Center, Strategic Laboratory, Unidade de Pronto Atendimento Dr Aloisio Muniz de Andrade, South America / Brazil / Sao Paulo / Presidente Prudente

Instituto Adolfo Lutz, Rapid Response Center, Strategic Laboratory, Servico de Atendimento de Especialidades de Itapevi, South America / Brazil / Sao Paulo / Itapevi

Arbovirology and Entomology, Bernhard Nocht Institute for Tropical Medicine, Arbovirology and Entomology, Bernhard Nocht Institute for Tropical Medicine, Africa / Gabon

Dallas County Health & Human Services Public Health Laboratory, DCHHS Sexual Health Clinic, North America / USA / Texas / Dallas

Antioquia, Laboratorio Departamental de Salud Publica de Antioquia, Antioquia, Laboratorio Departamental de Salud Publica de Antioquia, South America / Colombia / Antioquia

Laboratorio de Referencia Nacional de Viruas Inmunoprevenibles. Centro Nacional de Salud Publica. Instiuto Nacional de Salud, Laboratorio de Referencia Nacional de Viruas Inmunoprevenibles. Centro Nacional de Salud Publica. Instiuto Nacional de Salud, South America / Peru / Lima / Lima / Ate

Instituto Adolfo Lutz, Rapid Response Center, Strategic Laboratory, CTA Sao Mateus, South America / Brazil / Sao Paulo / Sao Paulo

Pathogen Genomic Laboratory, Institut National de Recherche Biomedicale (INRB), Pathogen Genomic Laboratory, Institut National de Recherche Biomedicale (INRB), Africa / Democratic Republic of the Congo / Mai-Ndombe / Mushie

Instituto Adolfo Lutz Strategic Laboratory, Fleury Medicina Dignóstica, South America / Brazil / Rio Grande do Sul / Porto Alegre

Instituto Adolfo Lutz, Rapid Response Center, Strategic Laboratory, Centro de Referencia e Treinamento DST AIDS, South America / Brazil / Sao Paulo / Sao Paulo

Instituto Adolfo Lutz, Rapid Response Center, Strategic Laboratory, Instituto Adolfo Lutz, Rapid Response Center, Strategic Laboratory, South America / Brazil / Sao Paulo / Sao Bernardo do Campo

Instituto Adolfo Lutz, Rapid Response Center, Strategic Laboratory, Centro de Prev e Assist Doencas Infecciosas Cepadi, South America / Brazil / Sao Paulo / Sao Caetano do Sul

Instituto de Salud Publica de Chile, Genetica Molecular and Subdepartamento de Virologia ISP Chile, South America / Chile / Region Metropolitana de Santiago / Vitacura

California Department of Public Health, California Department of Public Health, North America / USA / California

Centro de Desenvolvimento Científico e Tecnológico (CDCT), Centro Estadual de Vigilância em Saúde (CEVS) da Secretaria Estadual da Saúde (SES-RS), Centro de Desenvolvimento Científico e Tecnológico (CDCT), Centro Estadual de Vigilância em Saúde (CEVS) da Secretaria Estadual da Saúde (SES-RS), South America / Brazil / Rio Grande do Sul / Porto Alegre

Institute of Ecology and Evolution, University of Edinburgh, Nigeria Centre for Disease Control and Prevention, Africa / Nigeria / Borno

Instituto Adolfo Lutz, Rapid Response Center, Strategic Laboratory, Instituto Adolfo Lutz, Rapid Response Center, Strategic Laboratory, South America / Brazil / Sao Paulo / Santos

Los Angeles County Public Health Laboratories, Quest Diagnostic Nichols Institute, North America / USA / California

Department of Virology, National Institute of Health, Islamabad, Pakistan, Department of Virology, National Institute of Health, Islamabad, Pakistan, Asia / Pakistan / Punjab / Mandi Bahauddin

Laboratory of Respiratory Viruses and Measles, Oswaldo Cruz Institute, FIOCRUZ, Laboratory of Respiratory Viruses, Exanthematous, Enteroviruses and Viral Emergencies, South America / Brazil / Santa Catarina / Itajai

National Public Health Laboratory, National Centre for Infectious Diseases, National Public Health Laboratory, National Centre for Infectious Diseases, Asia / Singapore

Instituto Adolfo Lutz Strategic Laboratory, Instituto de Infectologia Emilio Ribas II Baixada Santista, South America / Brazil / Sao Paulo / Guaruja

Laboratory of Virology, National Institute of Allergy and Infectious Diseases (NIAID), National Institutes of Health (NIH), Laboratoire National de Santé Publique (LNSP), Africa / Republic of the Congo / Cuvette-Central

Charité Universitätsmedizin Berlin, Institut für Virologie, Charité Universitätsmedizin Berlin, Institut für Virologie/Labor Berlin, Europe / Germany / Berlin

Instituto Adolfo Lutz, Rapid Response Center, Strategic Laboratory, Servico de Atendimento Especializado de Sao Jose do Rio Preto, South America / Brazil / Sao Paulo / Sao Jose Do Rio Preto

Instituto Adolfo Lutz Strategic Laboratory, UBS II COHAB Presidente Prudente, South America / Brazil / Sao Paulo / Presidente Prudente

Department of Virology, Faculty of Medicine, University of Helsinki, Department of Virology, Faculty of Medicine, University of Helsinki, Europe / Finland

Laboratorio de Referencia Nacional de Viruas Inmunoprevenibles. Centro Nacional de Salud Publica. Instiuto Nacional de Salud, Laboratorio de Referencia Nacional de Viruas Inmunoprevenibles. Centro Nacional de Salud Publica. Instiuto Nacional de Salud, South America / Peru / Lima / Lima / San Luis

Thai Red Cross Emerging Infectious Diseases Clinical Center, King Chulalongkorn Memorial Hospital, Thai Red Cross Emerging Infectious Diseases Clinical Center, King Chulalongkorn Memorial Hospital, Asia / Thailand

Laboratory of Genomics and Bioinformatics, Comenius University Science Park, Public Health Authority of the Slovak Republic, Europe / Slovakia / Presov

Health Protection Agency, Porton Down, Health Protection Agency, Porton Down, Europe / United Kingdom

Research Institute for Tropical Medicine, San Lazaro Hospital, Asia / Philippines / National Capital Region

Instituto Adolfo Lutz Strategic Laboratory, Secrtetaria Municipal de Saude de Sertaozinho, South America / Brazil / Sao Paulo / Sertaozinho

Laboratory for Diagnostics of Zoonoses and WHO Centre, Institute of Microbiology and Immunology, Faculty of Medicine, University of Ljubljana, Laboratory for Diagnostics of Zoonoses and WHO Centre, Institute of Microbiology and Immunology, Faculty of Medicine, University of Ljubljana, Europe / Slovenia

Instituto Adolfo Lutz Strategic Laboratory, Centro de Referencia em AIDS SECRAIDS, South America / Brazil / Sao Paulo / Santos

Laboratorio de Referencia Nacional de Viruas Inmunoprevenibles. Centro Nacional de Salud Publica. Instiuto Nacional de Salud, Laboratorio de Referencia Nacional de Viruas Inmunoprevenibles. Centro Nacional de Salud Publica. Instiuto Nacional de Salud, South America / Peru / Lima / Lima / San Juan de Miraflores

Instituto Adolfo Lutz, Rapid Response Center, Strategic Laboratory, Hospital Cruzeiro do Sul, South America / Brazil / Sao Paulo / Osasco

Laboratorio de Referencia Nacional de Viruas Inmunoprevenibles. Centro Nacional de Salud Publica. Instiuto Nacional de Salud, Laboratorio de Referencia Nacional de Viruas Inmunoprevenibles. Centro Nacional de Salud Publica. Instiuto Nacional de Salud, South America / Peru / Lima / Lima / Rimac

Pathogen Genomic Laboratory, Institut National de Recherche Biomedicale, Pathogen Genomic Laboratory, Institut National de Recherche Biomedicale, Africa / Democratic Republic of the Congo / Tshopo

Instituto Adolfo Lutz Strategic Laboratory, Hospital Edmundo Vasconcelos, South America / Brazil / Sao Paulo / Sao Paulo

Instituto Adolfo Lutz, Rapid Response Center, Strategic Laboratory, Instituto Adolfo Lutz, Rapid Response Center, Strategic Laboratory, South America / Brazil / Sao Paulo / Campinas

Molecular Microbiology Laboratory, Department of Pathology, Molecular and Cell-Based Medicine, Icahn School of Medicine at Mount Sinai,, Molecular Microbiology Laboratory, Department of Pathology, Molecular and Cell-Based Medicine, Icahn School of Medicine at Mount Sinai,, North America / USA / New York

Public Health Ontario, Public Health Ontario, North America / Canada / Ontario

Institute of Ecology and Evolution, University of Edinburgh, Nigeria Centre for Disease Control and Prevention, Africa / Nigeria / Abia

División Diagnóstico Molecular Hospital México, División Diagnóstico Molecular Hospital México, North America / Costa Rica / San Jose / Coronado

Instituto Adolfo Lutz Strategic Laboratory, UPA 24H Brotas, South America / Brazil / Bahia / Salvador

Pathogen Genomic Laboratory, Institut National de Recherche Biomedicale (INRB), Pathogen Genomic Laboratory, Institut National de Recherche Biomedicale (INRB), Africa / Democratic Republic of the Congo / Sankuru

Laboratorio de Referencia Nacional de Viruas Inmunoprevenibles. Centro Nacional de Salud Publica. Instiuto Nacional de Salud, Laboratorio de Referencia Nacional de Viruas Inmunoprevenibles. Centro Nacional de Salud Publica. Instiuto Nacional de Salud, South America / Peru / Lambayeque / Chiclayo / Santa Rosa

Instituto de Salud Publica de Chile, Genetica Molecular and Subdepartamento de Virologia ISP Chile, South America / Chile / Valparaiso / Limache

Thai Red Cross Emerging Infectious Diseases Clinical Center and Faculty of Medicine, Chulalongkorn University, Suvarnabhumi Airport, Asia / Thailand

Instituto Adolfo Lutz Strategic Laboratory, UBS Jovaia, South America / Brazil / Sao Paulo / Guarulhos

Laboratorio de Referencia Nacional de Viruas Inmunoprevenibles. Centro Nacional de Salud Publica. Instiuto Nacional de Salud, Laboratorio de Referencia Nacional de Viruas Inmunoprevenibles. Centro Nacional de Salud Publica. Instiuto Nacional de Salud, South America / Peru / Arequipa / Arequipa / Mariano Melgar

Instituto de Salud Publica de Chile, Genetica Molecular and Subdepartamento de Virologia ISP Chile, South America / Chile / Region Metropolitana de Santiago / Quilicura

Instituto Adolfo Lutz Strategic Laboratory, USAFA Forte, South America / Brazil / Sao Paulo / Praia Grande

Laboratorio de Referencia Nacional de Virus Respiratorio. Centro Nacional de Salud Publica. Instituto Nacional de Salud., Laboratorio de Referencia Nacional de Virus Respiratorio. Centro Nacional de Salud Publica. Instituto Nacional de Salud., South America / Peru / La Libertad

Pathogen Genomic Laboratory, Institut National de Recherche Biomedicale (INRB), Pathogen Genomic Laboratory, Institut National de Recherche Biomedicale (INRB), Africa / Democratic Republic of the Congo / Kinshasa / Kinshasa

Instituto Adolfo Lutz Strategic Laboratory, UMS Parque Industrial Curitiba, South America / Brazil / Parana / Curitiba

Laboratorio de Referencia Nacional de Viruas Inmunoprevenibles. Centro Nacional de Salud Publica. Instiuto Nacional de Salud, Laboratorio de Referencia Nacional de Viruas Inmunoprevenibles. Centro Nacional de Salud Publica. Instiuto Nacional de Salud, South America / Peru / La Libertad / Trujillo / Laredo

Instituto Adolfo Lutz Strategic Laboratory, PR S da Familia Unidade de Saude Adalberto Rocha, South America / Brazil / Sao Paulo / Guarei

Institute of Ecology and Evolution, University of Edinburgh, Nigeria Centre for Disease Control and Prevention, Africa / Nigeria / Anambra

Institute for Hepatology,Shenzhen Third People's Hospital, Institute for Hepatology,Shenzhen Third People's Hospital, Asia / China / Shenzhen

Institute of Ecology and Evolution, University of Edinburgh, Nigeria Centre for Disease Control and Prevention, Africa / Nigeria / Gombe / Gombe

RIPHL at Rush University Medical Center, Quest Diagnostics, North America / USA / Illinois / Cook County / Chicago

Sicilian Regional Laboratory - AOUP "P. Giaccone" - University of Palermo, Sicilian Regional Laboratory - AOUP "P. Giaccone" - University of Palermo, Europe / Italy

Laboratorio de Referencia Nacional de Virus Inmunoprevenibles. Centro Nacional de Salud Publica. Instituto Nacional de Salud., Laboratorio de Referencia Nacional de Virus Inmunoprevenibles. Centro Nacional de Salud Publica. Instituto Nacional de Salud., South America / Peru / Ucayali

Institute of Ecology and Evolution, University of Edinburgh, Nigeria Centre for Disease Control and Prevention, Africa / Nigeria / Plateau

Public and Environmental Health Reference Laboratories, Pathology Queensland, Mater Pathology, Oceania / Australia / Queensland

Instituto Adolfo Lutz, Rapid Response Center, Strategic Laboratory, Hospital e Maternidade Sino Brasileiro, South America / Brazil / Sao Paulo / Osasco

Instituto Adolfo Lutz Strategic Laboratory, Secretaria Municipal de Saude Sao Carlos, South America / Brazil / Sao Paulo / Sao carlos

Instituto Costarricense de Investigacion y Enseñanza en Nutricion y Salud, Inciensa, Division de Microbiologia, Hospital Nacional de Niños Carlos Saenz Herrera, North America / Costa Rica / San Jose

Institute for Virology, Philipps-University Marburg, Institute for Virology, Philipps-University Marburg, Europe / Germany

Los Angeles County Public Health Laboratories, Quest Diagnostics Nichols Institute, North America / USA / California / Los Angeles County

Parasitology Laboratory, Parasitology Laboratory, Institute of Tropical Medicine of Sao Paulo, School of Medicine, University of Sao Paulo, South America / Brazil / Sao Paulo

Instituto Adolfo Lutz, Rapid Response Center, Strategic Laboratory, Instituto de Infectologia Emilio Ribas Sao Paulo, South America / Brazil / Sao Paulo / Sao Paulo

Osaka Metropolitan University, Graduate School of Medicine, Department of Virology and Parasitology, Osaka Metropolitan University, Graduate School of Medicine, Department of Virology and Parasitology, Asia / Japan / Tokyo

Laboratorio de Referencia Nacional de Viruas Inmunoprevenibles. Centro Nacional de Salud Publica. Instiuto Nacional de Salud, Laboratorio de Referencia Nacional de Viruas Inmunoprevenibles. Centro Nacional de Salud Publica. Instiuto Nacional de Salud, South America / Peru / Cusco / Cusco / Wanchaq

Instituto de Diagnostico y Referencia Epidemiologicos (INDRE), LESP Durango, North America / Mexico / Durango

Instituto Adolfo Lutz Strategic Laboratory, Secretaria Municipal de Saude de Feira de Santana, South America / Brazil / Bahia / Feira de Santana

RIPHL at Rush University Medical Center, ACL Laboratories, North America / USA / Illinois / Cook County / Chicago

Laboratorio de Referencia Nacional de Viruas Inmunoprevenibles. Centro Nacional de Salud Publica. Instiuto Nacional de Salud, Laboratorio de Referencia Nacional de Viruas Inmunoprevenibles. Centro Nacional de Salud Publica. Instiuto Nacional de Salud, South America / Peru / Loreto / Maynas / Iquitos

Instituto de Diagnostico y Referencia Epidemiologicos (INDRE), LESP Jalisco, North America / Mexico / Jalisco

Pathogen Genomics Lab, National Institute for Biomedical Research (INRB), Pathogen Genomics Lab, National Institute for Biomedical Research (INRB), Africa / Democratic Republic of the Congo / Sud-Ubangi

Instituto Adolfo Lutz Strategic Laboratory, Hosp. Municipla. Dr. Jose de Carvalho Florence, South America / Brazil / Sao Paulo / Sao Jose dos Campos

Laboratorio de Referencia Nacional de Viruas Inmunoprevenibles. Centro Nacional de Salud Publica. Instiuto Nacional de Salud, Laboratorio de Referencia Nacional de Viruas Inmunoprevenibles. Centro Nacional de Salud Publica. Instiuto Nacional de Salud, South America / Peru / Lima / Cañete / San Vicente de Cañete

Medical University of Vienna Center for Virology, Medical University of Vienna Center for Virology, Europe / Austria / Vorarlberg

Laboratorio de Referencia Nacional de Viruas Inmunoprevenibles. Centro Nacional de Salud Publica. Instiuto Nacional de Salud, Laboratorio de Referencia Nacional de Viruas Inmunoprevenibles. Centro Nacional de Salud Publica. Instiuto Nacional de Salud, South America / Peru / Ucayali / Coronel Portillo / Calleria

National Public Health Surveillance Laboratory (NPHSL), Santaros Klinikos, Lithuania, Europe / Lithuania / Vilnius

Instituto Adolfo Lutz, Rapid Response Center, Strategic Laboratory, Instituto Adolfo Lutz, Rapid Response Center, Strategic Laboratory, South America / Brazil / Sao Paulo / Sao Jose Do Rio Preto

National Institute of Health Research and Development, RS Mitra Keluarga Gading, Asia / Indonesia / DKI Jakarta / Jakarta Utara

Laboratorio de Referencia Nacional de Viruas Inmunoprevenibles. Centro Nacional de Salud Publica. Instiuto Nacional de Salud, Laboratorio de Referencia Nacional de Viruas Inmunoprevenibles. Centro Nacional de Salud Publica. Instiuto Nacional de Salud, South America / Peru / La Libertad

Instituto Adolfo Lutz, Rapid Response Center, Strategic Laboratory, Vigilancia Epidemiologica de Sao Bernardo do Campo, South America / Brazil / Sao Paulo / Sao Bernardo Do Campo

Laboratoire National de Reference, Institut National de Sante Publique of Burundi, Laboratoire National de Reference, Institut National de Sante Publique of Burundi, Africa / Burundi / Bujumbura-Nord

Instituto de Salud Publica de Chile, Genetica Molecular and Subdepartamento de Virologia ISP Chile, South America / Chile / Valparaiso / Villa Alemana
